# Multi-trait GWAS of atherosclerosis detects novel pleiotropic loci

**DOI:** 10.1101/2021.05.21.21257493

**Authors:** Tiffany R. Bellomo, William P. Bone, Brian Y. Chen, Katerina A. B. Gawronski, David Zhang, Joseph Park, Michael Levin, Noah Tsao, Derek Klarin, Julie Lynch, Themistocles L. Assimes, J. Michael Gaziano, Peter W. Wilson, Kelly Cho, Marijana Vujkovic, the VA Million Veteran Program, Christopher J. O’Donnell, Kyong-Mi Chang, Phil S. Tsao, Daniel J. Rader, Marylyn D. Ritchie, Benjamin F. Voight, Scott M. Damrauer

## Abstract

**Rationale:** Although affecting different arterial territories, the related atherosclerotic vascular diseases coronary artery disease (CAD) and peripheral artery disease (PAD) share similar risk factors and have shared pathobiology. Analysis of their shared genetic architecture, along with that of common risk factors, may identify novel common biology.

**Objective:** To identify novel pleiotropic genetic loci associated with atherosclerosis and provide a better understanding of biological pathways underlying atherosclerosis.

**Methods and Results:** Summary statistics from genome wide association studies (GWAS) of nine known atherosclerotic (CAD, PAD) or atherosclerosis risk factors (body mass index, smoking initiation, type 2 diabetes, low density lipoprotein (LDL), high density lipoprotein, total cholesterol, and triglycerides) were combined to perform 15 separate multi-trait genetic association scans which resulted in 31 unique novel pleiotropic loci not yet reported as genome-wide significant for their respective traits. Colocalization with single-tissue eQTLs identified 34 candidate causal genes across 14 of the detected signals. Notably, the signal between PAD and CAD at the *VDAC2* locus (rs7088974) colocalized with *VDAC2* expression in aorta and tibial artery tissues. Additionally, the signal between PAD and LDL at the *PCSK6* locus (rs1531817) affects *PCSK6* splicing in human liver tissue and induced pluripotent derived hepatocyte like cells.

**Conclusions:** Joint analysis of related atherosclerotic disease traits and their risk factors allowed identification of unified biology that may offer the opportunity for therapeutic manipulation. *VDAC2* and *PCSK6* represent possible shared causal biology where existing inhibitors may be able to be leveraged for novel therapies.

## INTRODUCTION

Atherosclerotic vascular disease is a leading cause of cause of death worldwide^1-3^ and can affect multiple arterial territories. Although clear differences in disease pathobiology exist^4^, epidemiological analyses have shown both coronary artery disease (CAD) and peripheral artery disease (PAD) share similar risk factors and frequently co-occur in the same patients^5-7^. These risk factors include dyslipidemia, obesity, and other environmental factors^8^. PAD patients with concomitant CAD are known to experience more extensive and aggressive disease^9^.

The genetics of CAD have been well characterized and a number of genome-wide association studies (GWAS) have identified over 200 genetic risk loci with robust connections to CAD^10-12^. For most loci, however, underlying mechanisms by which these loci influence CAD risk remains unclear. Although PAD has been less intensively studied, recent work has identified 21 total risk loci associated with PAD risk^6,13^. Genetic correlation studies have demonstrated a high degree of shared genetic architecture between CAD and PAD (rg 0.67)^14^. This genetic correlation, based on shared pathobiology, can be leveraged to identify novel pleiotropic genetic architecture common to both disease traits^15,16^.

The development of statistical approaches for multi-trait GWAS meta-analysis has facilitated joint analyses of traits with substantial evidence for a common pathophysiological basis to elucidate shared genetic etiology^6^. Furthermore, correlated causal risk factors can also be included in these multi-trait GWAS analyses to provide insight on their shared genetic pathways^16-20^. Although previous studies have analyzed CAD jointly with a second trait to understand shared genetic etiology^16,17^, there have been no studies which evaluate atherosclerosis endpoints jointly with multiple cardiometabolic causal risk factors.

In this study, we performed a series of N-weighted multivariate genome-wide-association meta-analyses (N-GWAMA)^15^ using different combinations of nine atherosclerotic or atherosclerosis risk factor traits, and identified 31 unique pleiotropic loci not previously associated with any analyzed trait combination. We subsequently used single-tissue expression quantitative trait loci (eQTL) colocalization analysis at these loci to identify 34 suggestive candidate causal genes and their tissue site of action. Some of these causal gene candidates provide potential opportunities for drug repurposing to treat atherosclerotic vascular disease, including *VDAC2* and *PCSK6*. Ultimately, this study provides a better understanding of biological pathways underlying atherosclerosis to inform future therapeutic development.

## METHODS

This study was approved by the U.S. Department of Veterans Affairs Central Institutional Review Board and the University of Pennsylvania Institutional Review Board. All participants gave written informed consent for study participation.

### Genetic Association Data

We collected the summary statistics from the largest published GWAS to maximize our power for novel discovery. PAD summary statistics were taken from the recent VA Million Veteran Program analysis with 31,307 PAD cases and 211,753 PAD controls^6^ and can be accessed in dbGAP (phs001672.v2.p1). CAD data were taken from CARDIoGRAMplusC4D combined with the UK BioBank (UKBB)^10^ and consisted of 122,733 CAD cases and 424,528 CAD controls and can be accessed through Mendeley doi:10.17632/gbbsrpx6bs.1. Data for body mass index (BMI) (meta-analysis of GIANT and UKBB; 806,834 individuals; ^21^), type 2 diabetes (T2D) (meta-analysis of consortia; 228,499 cases and 1,178,783 controls; ^22^), smoking initiation (SMK) (UKBB ; 462,690 individuals; ^23^), and 4 lipids traits (meta-analysis of MVP and GLGC data; 723,000; ^24^) were obtained from the public domain as well (**Supplementary Table 1**). We confirmed the shared genetic etiology of PAD and CAD, as well as 7 of their shared risk factors (BMI, SMK, T2D, low density lipoprotein (LDL), high density lipoprotein (HDL), total cholesterol (TC), and triglycerides(TG)) by applying cross-trait LD score regression (**Supplementary Table 2 and Supplementary Figure S1**).

### N-GWAMA Multi-trait GWAS

Using the summary statistics from publicly available single-trait GWAS (**Supplementary Table 1**), we performed 15 N-GWAMA^15^ multi-trait GWAS described in supplemental methods centered around PAD, CAD, and the following atherosclerotic risk factor traits: BMI, T2D, SMK, LDL, HDL, TC, and TG. We first performed a bivariate GWAS for PAD and CAD followed by a series of trivariate GWAS combining PAD, CAD, and one of 7 traits correlated traits that represented traditional atherosclerotic risk factors. We also performed a series of bivariate GWAS between PAD and these seven traits individually, given that a series of bivariate GWAS between CAD and most of these seven traits has already been performed^17^.

Each N-GWAMA multi-trait GWAS resulted in a set of independent loci represented by a lead variant. We defined an independent locus as the genomic region that includes all variants within 1 MB of the lead variant and any other variants that were in LD of r^2^ > 0.2 with the lead variant using the 1000 Genomes European ancestry cohort (1kG EUR)^14^. We applied a series of filters to the independent loci detected in the N-GWAMA multi-trait GWAS to remove loci that were not plausibly pleiotropic or did not represent novel associations.

To report the locus as novel, we required that the lead variant was nominally associated (p<5×10^−3^) with all the individual traits involved in the multi-trait analysis. We also required that none of the variants at an independent locus (SNPs within 1MB or LD of r^2^ > 0.2 of the lead variant) were previously associated with any of the traits used in the N-GWAMA multi-trait GWAS^25^ (Figure 1). Further detail on this pipeline is available in the supplement and the code is available on github at https://github.com/Bellomot/Athero_NGWAMA_Multitrait_GWAS or from the authors upon request (B.F.V, S.M.D.). We also excluded loci in the HLA region from these experiments due to the difficulty of interpreting the independent signals of these loci.

**Figure 1.**
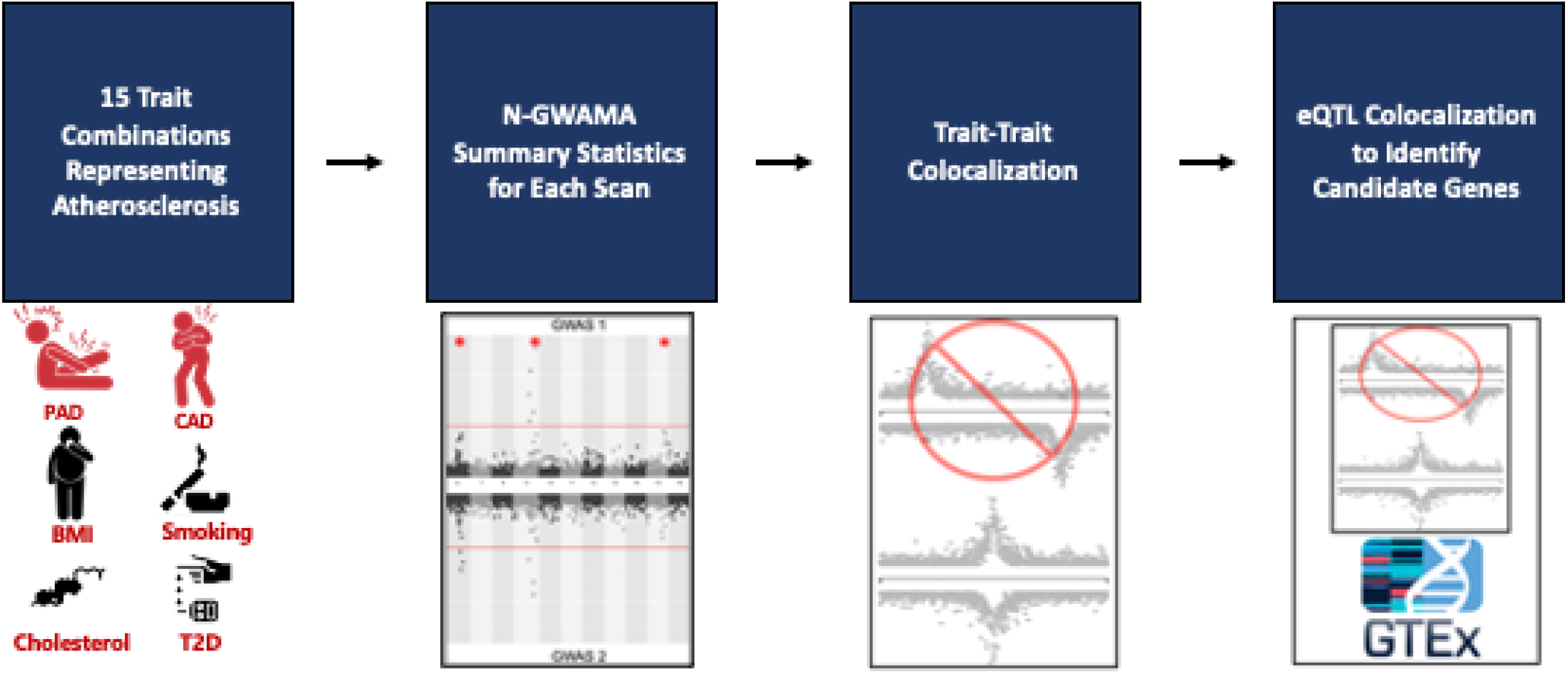
Flowchart of multi-trait analysis and candidate gene results. 9 traits were analyzed in 15 different bivariate and trivariate scans that best represented atherosclerosis. The summary statistics from all scans were filtered by single trait P values and loci within 500kb or in LD (EUR r^2^⍰>⍰0.2 1kG EUR) with the known trait being tested according to the GWAS catalog, resulting in 150 unique loci. Trait to trait colocalization with a threshold of a conditional posterior probability of colocalization > 0.8 was performed to ensure evidence of a shared causal SNP between each trait. The resulting 31 unique loci were run through single tissue eQTL analysis using GTEx v8 to identify candidate causal genes and tissues for each locus. 34 unique genes were identified among 14 loci.

We implemented multiple testing correction given that we performed 15 multi-trait GWAS for combinations of related traits. Due to the high correlation between each of the multi-trait GWAS we performed, a Bonferroni correction (p<3×10^−9^) for each trait combination test would be too stringent. Thus, q-values were calculated by concatenating all of the SNP results from the 15 N-GWAMA multi-trait GWAS using the R package qvalue. We defined nominal genome-wide significance as having both a genome-wide significant (p<5×10^−8^) N-GWAMA multi-trait P-value and multi-trait N-GWAMA q-value < 5×10^−5^. We also used a null distribution Z-score sampling strategy to estimate an α = 0.05 P-value threshold for this set of N-GWAMA multi-trait GWAS. We first drew 10,000 sets of 1 million samples from a 15-dimensional multivariate normal distribution, which estimates the distribution of the null Z-scores of the 15 N-GWAMA multi-trait GWAS. The 15-dimensional multivariate normal distribution was centered at the origin, and we used the estimated correlation between the Z-scores of the 15 N-GWAMA multi-trait GWAS as the correlation matrix (**Supplementary Table 3**). We kept the most extreme Z-score from each of the 10,000 sets and then identified the 95^th^ percentile of the most extreme Z-scores as our α = 0.05 threshold. We defined experiment wide significance as the 95^th^ percentile Z-score of 5.87, which corresponds to a P-value of 4.3×10^−9^.

### Trait-trait Colocalization

For each multi-trait GWAS, we assessed the evidence of a shared causal variant at each significant locus by performing colocalization analysis between the trait signals using COLOC for bivariate GWAS and MOLOC for trivariate GWAS^26,27^ (**Supplementary Table 4**). For this analysis, we applied a 500 KB window (+/- 250 KB) around the lead variant. A conditional probability of colocalization (e.g., PP4/ (PP3 + PP4)) is defined as the posterior probability of colocalization conditioned on the presence of a signal for each trait. A probability of ≥ 0.8 was considered significant. Loci that had a conditional probability of colocalization > 0.5 and < 0.8 were visually inspected using LocusZoom plots (**Supplementary Table 5**). If the LD structure suggested additional associations unlinked to the leading variant in the region, approximate conditional analysis was performed (see Approximate Conditional Analysis below).

### Single-Tissue Gene Expression Colocalization

We performed single-tissue colocalization analysis to prioritize candidate causal genes implicated in each N-GWAMA multi-trait GWAS using RNA-seq data obtained from the Genotype-Tissue Expression (GTEx) project^26^. Using the signif_variant_gene_pairs.txt.gz GTEx v8 files, we identified the list of genes and tissues for which each N-GWAMA lead variant was a significant single-tissue expression quantitative trait locus (eQTL) in GTEx v8 (**Supplementary Table 6**). We then performed colocalization between either CAD or PAD, as determined by which trait had the most significant lead SNP at each locus, and each single-tissue eQTL signal from GTEx v8^26^. The window of colocalization was 500 KB spanning the lead variant. Similar to trait-trait colocalization analysis, our threshold to classify the traits as colocalized was a conditional probability of colocalization (PP4/ (PP3 + PP4)) ≥ 0.8. We visually inspected LocusZoom plots for loci where colocalization analysis resulted in a conditional probability of colocalization < 0.8 but > 0.5 and performed approximate conditional analysis when the LD structure suggested possible allelic heterogeneity.

### Approximate Conditional Analysis

For each locus that showed evidence of multiple independent signals, we performed approximate conditional analysis on variants that appeared to be associated with the trait of interest independently of the lead variant (**Supplementary Table 7**). This analysis was necessary given that the presence of multiple associated variants in a region violates the assumptions of COLOC^26^. Potential nearby association signals were identified using LocusZoom plots and the LDassoc tool of LDlink^28^. We performed approximate conditional analysis using GCTA-COJO with 1000 Genome Project data (European samples, n=503) as a reference panel^29,30^. We conditioned the lead variant on the most associated variant for each potential confounding signal identified at the locus. We then repeated the colocalization experiment on the locus using the conditional variant P-values. A full list of traits, the lead variants, and the conditioned variants for each conditional analysis are provided in the supplement (**Supplementary Table 7**).

### Splicing Quantitative Trait Locus Colocalization

We performed a colocalization analysis between the PAD signal at the *PCSK6* locus and the GTEx v8 liver tissue splicing quantitative trait locus (sQTL) signal with the intron ID: 101365044:101366196:clu_14775 from the Liver.v8.sqtl_allpairs.txt.gz. We also identified this intron signal in the PHLiPs HLC sQTL data by lifting over the start and stop of this intron to hg19 (101905249:101906401)^31^. We then performed colocalization analysis between the HLC sQTL signal and the PAD signal as well as the HLC sQTL signal and the GTEx v8 liver tissue sQTL signal (**Figure 3**).

### Allele Specific Expression

We assessed whether allele specific expression (ASE) was a potential mechanism of the liver sQTL at the *PCSK6* locus. To perform this analysis, we first identified six coding variants or UTR3 variants in *PCSK6* that would be informative of whether a *PCSK6* isoform included or excluded exon 14 (GRCh38 chr15:101364999-101365044 of the *PCSK6* isoform CCDS73793 with ID ENST00000611967.4) (**Supplementary Table 8**). We then identified individuals in the GTEx allele expression analyses who had liver tissue eQTL data, were heterozygous for rs1531817, and were heterozygous for one of the six exon 14 informative variants^32^. For each sample identified, we performed a binomial test to assess if the ratio of reference to alternate alleles deviated from the expected null ratio of approximately 0.5 for all these variants. Further detail on the GTEx allele expression analyses can be found in the original publication^32^.

## RESULTS

### Multi-trait GWAS analysis results

We performed 15 N-GWAMA scans to detect novel loci not previously reported as genome-wide significant for any of their respective traits (**Supplementary Table 9**). A total of 150 loci were multi-variate genome-wide significant with all single trait P-values between < 5×10^−3^ and > 5×10^−8^ (**Supplementary Table 4**). A total of 31 unique loci were nominal genome-wide significant, met our colocalization criteria, and thus represent novel pleotropic loci not previously reported in a single trait GWAS for their respective multi-trait GWAS (**Table 1, Supplementary Figures S2-S34**). 14 of the nominal genome-wide significant loci colocalized with one or more single-tissue eQTLs representing 34 different genes across 46 different tissues (**Supplementary Table 6**). Finally, five loci colocalized with eQTLs for genes that have been implicated in atherosclerosis by previous studies.

### VDAC2 and apoptosis

We detected a nominal genome-wide significant signal with PAD and CAD (bivariate P=7.8×10^−9^) at the *VDAC2* locus rs7088974 (**Table 1**). Variants at this locus have been found to be associated with body fat percentage, waist circumference, and BMI in previous GWAS in the open GWAS project (Supplementary Table 5). Although variants near this locus have also been associated with smoking behavior, our results suggest that the locus we detected is independent of smoking behavior^33^ (**Supplementary Figure S35**). This signal colocalized with *VDAC2* mRNA expression in multiple vascular tissues relevant to atherosclerosis, including aorta and tibial artery (Figure 2 and Supplementary Table 6). The direction of effect at this locus was concordant for PAD, CAD, and all gene tissue pairs: the allele that associated with increased *VDAC2* expression (EA = C, EAF = 0.57) is associated with increased PAD and CAD (β=-0.14, SE=0.02, P=7.7×10^−9^).

**Table 1.**
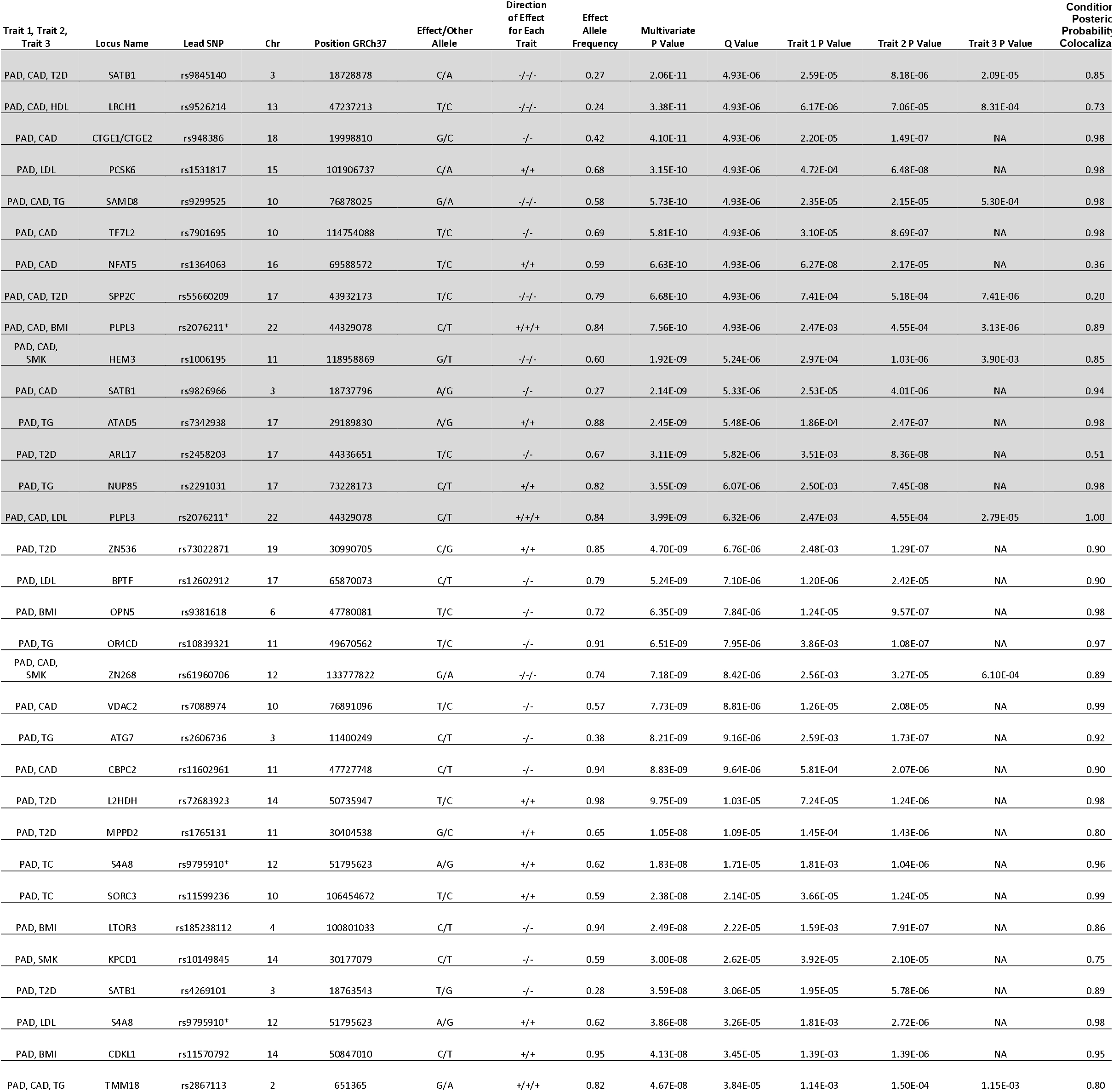
Atherosclerosis trait NGWAMA analysis and results. Trait 3 P Value will have a value of NA if there were only 2 traits analyzed. The Q value was calculated based on all of the SNP results from the 15 N-GWAMA multi-trait GWAS. Conditional posterior probability represents the PP4/(PP3+PP4) probability of the trait to trait colocalization analysis. * indicates that the Lead SP was detected in another trait combination scan. Loci in gray met the experiment-wide significance threshold (P-value < 4.3×10-9) Abbreviations: PAD/peripheral artery disease; CAD/coronary artery disease; BMI/body mass index; T2D/type 2 diabetes; SMK/smoking; HDL/high density lipoprotein; LDL/low density lipoprotein; TG/triglycerides; TC/total cholesterol; Chr/chromosome.

**Figure 2.**
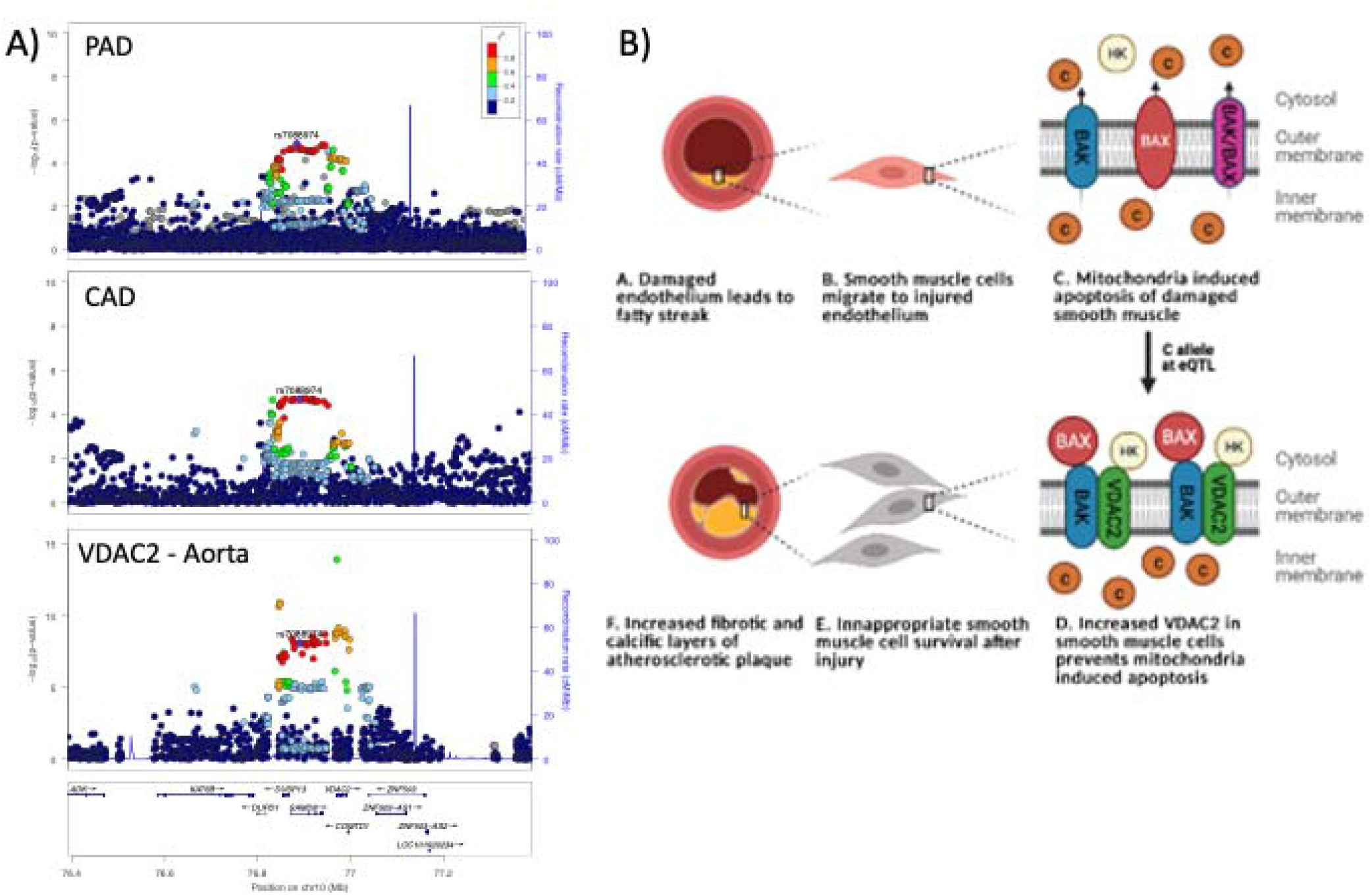
VDAC2 locus with a lead SNP of rs7088974. A) Pleiotropic signal between PAD and CAD with an eQTL for VDAC2 in aortic artery tissue from GTEx v8. B) Hypothesized mechanism for the role of VDAC2 in atherosclerosis. Damaged endothelium leads to smooth muscle cell migration and if a person has a C allele at this locus, VDAC2 is overexpressed. Increased VDAC2 allows smooth muscle cells to become resistant to mitochondrial induced apoptosis and this inappropriate cell survival causes plaque progression.

### PCSK6 and lipid metabolism

We detected an experiment-wide significant signal with PAD and LDL (bivariate P=3.2×10^−10^) at the *PCSK6* locus. A rare coding variant in this region has been reported to associate with LDL^34^; however, the coding variant (NP_002561.1:p.Thr964Met, rs34631529) and our lead SNP (rs1531817) are not in linkage disequilibrium (r^2^ = 0.0086 1kG EUR) based on data from the 1000 Genomes Project^14^, indicating that we detected a different signal at this locus. To further differentiate whether our signal was novel, we performed an additional conditional analysis on the coding variant rs34631529 in PAD data without any notable changes in the *PCSK6* locus signal (**Supplementary Figure S36**). Previous GWAS have found that variants at this locus are associated with inflammatory markers^35-40^.

To better understand how genetic variation at the *PCSK6* locus influences circulating lipid levels, we investigated the association of the bivariate lead SNP at this locus in the publicly available GWAS of NMR lipid subfractions: extra-small subfrations (XS), extra-large subfractions (XL), HDL, intermediate density lipoprotein (IDL), LDL, and very-low density lipoprotein (vLDL)^41^. We found our lead SNP (rs1531817) had a nominal positive association with medium VLDL particles (β=0.03, SE=0.01, P=9×10^−3^), total lipids in medium VLDL (β=0.03, SE=0.01, P=0.02), TG in medium VLDL (β=0.02, SE=0.01, P=0.03), serum TG (β=0.02, SE=0.01, P=0.03), and TG in large VLDL (β=0.02, SE=0.01, P=0.03).

Given the association of our lead variant with increased lipid subfractions, we considered the potential mechanisms that would produce a gain-of-function effect consistent with our effect allele. Our lead variant was a sQTL for *PCSK6* in liver tissue in GTEx v8 liver tissue (**Figure 3**). To identify a potential experimental model of this splicing change, we searched for this sQTL in the Phenotyping Lipid traits in iPS derived hepatocytes Study (PhLiPS Study)^42^. The signal at *PCSK6* colocalized with an sQTL in these data as well (**Figure 3**).

**Figure 3.**
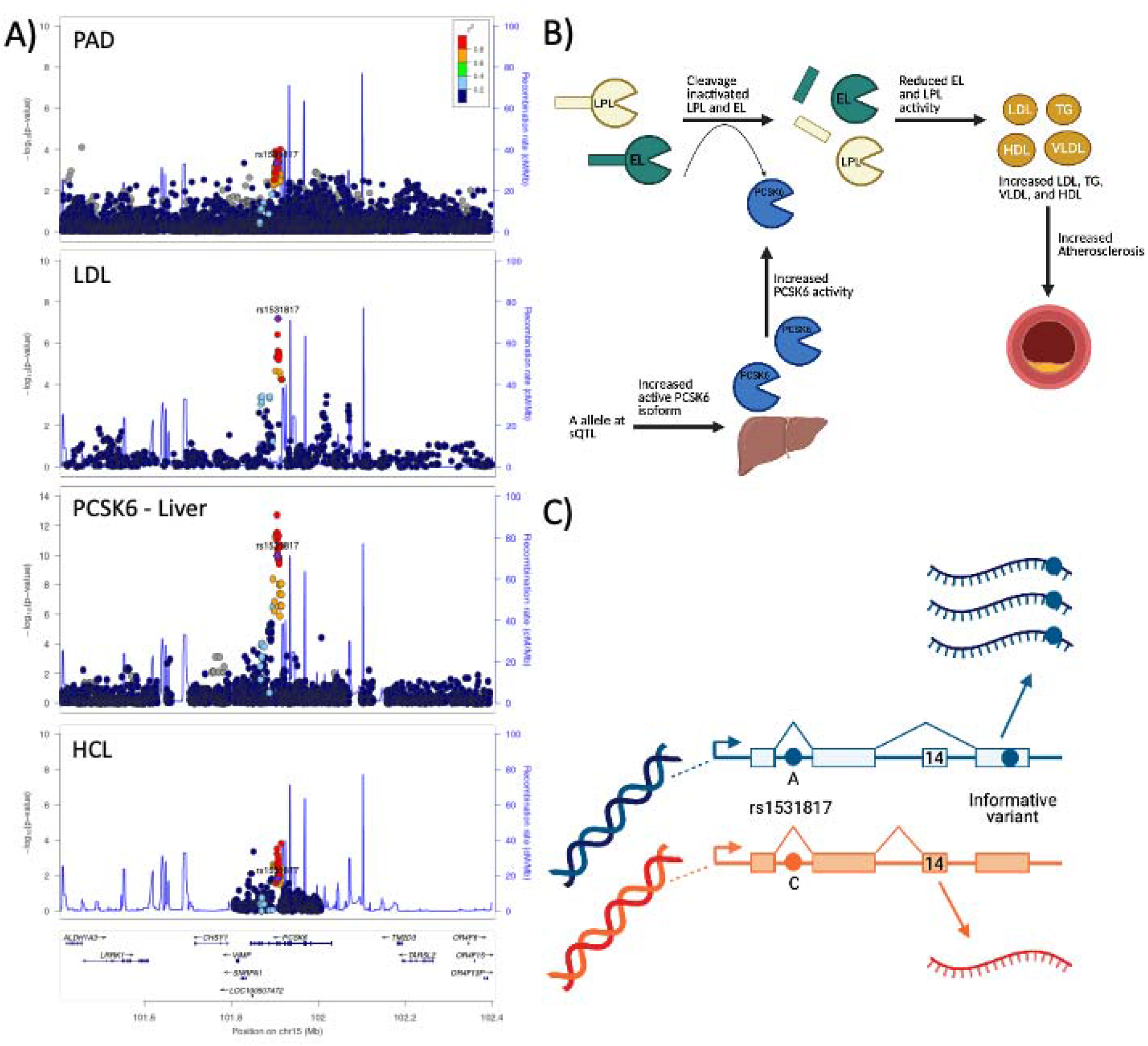
PCSK6 locus with a lead SNP of rs1531817. A) Pleiotropic signal between PAD and LDL with an sQTL for PCSK6 in liver tissue. This locus also colocalized with Hepatocyte-like cells (HLCs) in vitro. B) Hypothesized mechanism for the effects of PCSK6 on atherosclerosis. If a person has an A allele at the PCSK6 locus, exon 14 will be spliced out of PCSK6, which ultimately leads to increased excretion of the active isoform of PCSK6. Active PCSK6 will cleave and inactivate endothelial lipase (EL) and lipoprotein lipase (LPL) which will allow lipid levels of LDL, TG, vLDL, and HDL to increase and worsen atherosclerotic burden. C) Allele specific expression experiments. Six individuals in the liver tissue of the GTEx allelic expression data were heterozygous for our lead SNP of interest and one of six exon 14 informative variants. Allele 1 (shown in blue) has the A allele of our lead SNP of interest, which results in inclusion of exon 14 and increased activity of the transcript relative to allele 2 (shown in orange). The allelic origin of the transcript was assessed by the ratio of reads that had the reference or alternative allele and then was tested for significant deviation from the expected null ratio.

We assessed the evidence of ASE at the *PCSK6* locus in the liver tissue of the GTEx allelic expression data^32^ (**Figure 3**). There were six individuals with liver tissue data available who were heterozygous for both the sQTL lead SNP, rs1531817, and one of six exon 14 informative variants. For each of these individuals, we performed a binomial test to assess if the ratio of reads that had the reference or alternate allele of the informative variant deviated from the estimated null ratio (Supplementary Table 8). Two of these six binomial tests were nominally significant (P < 0.05).

### Other candidate indicate genes with known atherosclerotic biology

We detected a trivariate GWAS signal with PAD, CAD, and SMK (trivariate P=1.9×10^−9^) at the *HEM3* locus rs1006195 (**Table 1**). This variant was genome-wide significant in previous GWAS studies for several cardiometabolic traits including Apolipoprotein A1 levels, waist-hip ratio, BMI, fat mass percentage, HDL, and T2D^10,35,43-45^. This pleiotropic signal colocalized with *HMBS* and *VPS11* mRNA expression in several tissues (**Supplementary Figure S14 and Supplementary Table 6**). *HMBS* (β=0.28, SE=0.03, P=4.1×10^−23^) demonstrated the opposite direction of effect to atherosclerosis and SMK, while *VPS11* (β=-0.25, SE=0.03, P=6.0×10^−15^) demonstrated the same direction of effect in all tissue pairs.

We detected a nominal genome-wide significant signal with PAD and TC (bivariate P=2.4×10^−8^) at the *SORCS3* locus rs11599236 (**Table 1**). This locus was previously found to be genome-wide significant in GWAS studies of mood disorders in the open GWAS project (**Supplementary Table 5**) and this signal colocalized with *SORCS3* mRNA expression in pituitary tissue (**Supplementary Figure S34**). The same direction of effect was noted for both traits and the gene tissue pair increased *SORCS3* associated with increased PAD and TC (β=0.31, SE=0.05, P=5.5×10^−8^).

Finally, we detected a trivariate signal with PAD, CAD, and HDL (trivariate P=3.4×10^−11^) at the *LRCH1* locus rs9526214 (**Table 1**). This locus had evidence of allelic heterogeneity when we reviewed the regional association plots, which led us to perform approximate conditional analyses on the pleiotropic signal lead SNP rs9316223 and the resulting conditional probability of colocalization met our criteria (Supplementary Table 6). This locus has been found to be genome-wide significant in previous GWAS studies for platelets, systolic blood pressure, and stroke in the open GWAS project (**Supplementary Table 5**). This signal colocalized with *LRCH1* mRNA expression in tibial artery, whole blood, and other tissues (**Supplementary Figure S12 and Supplementary Table 6**). The same direction of effect was noted for all three traits and the gene tissue pair: increased *LRCH1* associated with increased PAD, CAD, and HDL (β=-0.09, SE=0.02, P=1.3×10^−8^).

## DISCUSSION

To advance our understanding of the genetic etiology of atherosclerosis, different combinations of nine known atherosclerotic or atherosclerosis risk factor traits were used to perform 15 N-GWAMA scans which resulted in 31 unique novel pleiotropic loci not yet reported as genome-wide significant for their respective traits (**Figure 1**), and one novel signal at a recently reported locus associated with one trait. Colocalization with single-tissue eQTLs identified 3 candidate causal genes across 14 of the detected signals. Five of these loci had candidate causal genes previously associated with atherosclerosis through other studies.

### VDAC2 increases atherosclerosis through inappropriate smooth muscle cell survival

We identified a novel pleiotropic association at the *VDAC2* locus between PAD and CAD. Colocalization experiments support *VDAC2* as the causal gene at this locus. *VDAC2* is a mitochondrial porin family protein that transports small metabolites and other ions across the outer mitochondrial membrane^46^. Although *VDAC2* has been well studied for its role in atrial cell cardiac contractility^46,47^ and interaction with endothelium-dependent nitric oxide synthase to worsen pulmonary hypertension^48^, it has also been reported to increase inappropriate cell survival after vascular injury^49^. Repeated cycles of vascular endothelial and smooth muscle cell damage with subsequent repair contribute to the buildup of extracellular lipid deposits that cause occlusive vascular lesions, especially when defects in apoptotic regulation of these cells exist^50^. *VDAC2* creates a complex with the integral mitochondrial membrane protein BAK, thereby inhibiting BAK binding domains and BAK dependent mitochondrial apoptosis^51^. Consistent with these observations, *VDAC2* was found to be significantly elevated in human atherosclerotic lesion cultures from surgical revascularization plaque samples^49^. We hypothesize that the causal variant at this locus leads to *VDAC2* overexpression and inhibition of mitochondrial dependent apoptosis^49^ in smooth muscle cells that have migrated to injured endothelium in atherosclerotic plaques (**Figure 2**). This process allows for repeated cycles of vascular cell damage, subsequent inappropriate repair, and plaque progression. We observed that *VDAC2* was associated with increased PAD and CAD, suggesting that the absence of *VDAC2* potentially allows for smooth muscle cell apoptosis and plaque resolution.

Multiple inhibitors of *VDAC2* have been reported in the literature and may offer the opportunity for therapeutic repositioning or additional pharmaceutical development. The angiotensin receptor blocker telmisartan has been shown to significantly downregulate *VDAC2* expression and other apoptotic regulators in human umbilical artery endothelial cells, suggesting this mechanism contributes to telmisartan’s antiatherogenic and plaque stabilizing properties^52^. Another non-specific *VDAC2* inhibitor hydroxysafflor yellow A (HSYA), a traditional medication, is approved in China to reduce LDL induced cellular injury^53^. Additionally, efsevin, a dihydropyrrole carboxylic ester compound, and erastin analog A9, an inducer of iron dependent cell death (ferroptosis), bind directly to *VDAC2* and inhibit *VDAC2* activity ^54-56^.

### PCSK6 activity effects lipid levels and smooth muscle cell migration

We identified a signal at the *PCSK6* locus that has a bivariate association with PAD and LDL and provide strong evidence in support of *PCSK6* as the causal gene at the *PCSK6* locus. *PCSK6* is a calcium-dependent serine endoprotease that cleaves proteins to active and inactive forms depending on the target protein^57^. This enzyme has been studied in the context of atherosclerosis influencing both smooth muscle cell migration and lipid profiles. There is convincing experimental evidence to suggest smooth muscle cell migration in injured arteries is facilitated by cytokine induced *PCSK6* expression that activates matrix metalloproteinases (MMP14/MMP2)^58^. This smooth muscle cell mechanism may explain the association of *PCSK6* with carotid intima-media thickness in a candidate gene study^59^. In that study, the lead variant associated with maximum progression of carotid intima-media thickness and was the same as the lead variant identified in our bivariate scan between LDL and PAD.

Our data support the genetic colocalization of *PCSK6* gene expression and LDL levels. This suggests *PCSK6* also influences lipid metabolism, a known upstream cause of cause atherosclerotic progression (**Figure 3**). This is in agreement with the known role of *PCSK6* in cleavage and inactivation of endothelial lipase (EL) and lipoprotein lipase (LPL)^60^. Reduced EL and LPL activity can both lead to hyperlipidemia^61^ (**Figure 3**). This hypothesized mechanism is supported by the recently reported rare coding variant in *PCSK6* associated with decreased LDL, possibly due to decreased activity of *PCSK6*, increased activity of EL and LPL, and ultimately decreased LDL^34^. We found the lead variant rs1531817 was nominally positively associated with circulating lipoprotein subfraction levels as measured by NMR spectroscopy, suggesting this locus may influence atherosclerosis progression through lipid metabolism. In our analyses, the A allele at our lead variant associates with the exclusion of exon 14 via RNA splicing of *PCSK6* in liver tissue^62,63^ (see methods above), which results in an active *PCSK6* isoform^64^ (**Figure 3**). We hypothesize this isoform would lead to overall increased activity of *PCSK6* in systemic circulation and increased lipid fractions. There was also nominal evidence of ASE at this locus in two of the six GTEx saples with informative *PCSK6* variants. It remains to be determined if changes in *PCSK6* activity result in altered lipid metabolism and therefore has downstream effects on PAD or if altered *PCSK6* activity effects LDL and PAD separately through independent mechanisms. In all likelihood, it is a combination of LDL dependent and independent mechanisms that link *PCSK6* to PAD.

There are several non-FDA approved, non-specific *PCSK6* inhibitors that influence lipid metabolism: both alpha1-antitrypsin Portland (alpha1-PDX)^65^ and profurin^66^ inhibit and partially inhibit *PCSK6* respectively to prevent EL cleavage and modify cholesterol transport. Other inhibitors of *PCSK6* are used for other disease processes and have the potential to be repurposed for atherosclerotic disease: Pf-pep is a positively charged pentamolecule that is a competitive inhibitor of *PCSK6* used for cartilage breakdown in osteoarthritis^67^ and dicoumarols (DC), specifically DC2, are small molecules that non-specifically competitively inhibit rat *PCSK6* to prevent virus propagation^68^.

### Limitations

We acknowledge there are several limitations to this study. First, there is sample overlap between several of our single trait summary statistics files. The N-GWAMA method attempts to account for this, but if the correction for the overlap was insufficient this could inflate our false discovery rate. Second, we selected nine atherosclerotic and cardiometabolic traits based on conventional relationships with atherosclerosis; however, there are likely multifactorial and multidirectional relationships within this group of traits. It is possible that some of the novel loci represent the interaction between traits instead of the intended representation of atherosclerosis as we have interpreted it. It is also possible that including other sets of cardiometabolic risk factors may identify additional novel loci.

## CONCLUSIONS

We have shown that publicly available GWAS data can be leveraged to perform multi-trait scans with N-GWAMA methods to identify novel loci that unify atherosclerosis. In this study, 31 unique nominal genome-wide significant loci were associated jointly with PAD and other atherosclerotic traits. These loci may represent novel genetic etiologies of atherosclerosis. A total of 34 candidate causal genes were identified across 14 novel pleiotropic loci and among those, *VDAC2* and *PCSK6* represent possible causal biology with known inhibitors that have large potential to be therapeutic targets for atherosclerosis. These results highlight the biological underpinnings of atherosclerosis and the potential to develop non-invasive medical treatments for atherosclerosis.

## Data Availability

The full summary level association data from the MVP trans-ancestry PAD meta-analysis from this report are available through dbGAP, accession code phs001672. Additional data that support the findings of this study are available on request from the corresponding author (S.M.D.); these data are not publicly available due to U.S. Government and Department of Veteran's Affairs restrictions relating to participant privacy and consent. Data contributed by CARDIoGRAMplusC4D investigators are available online (http://www.CARDIOGRAMPLUSC4D.org/). The genetic and phenotypic UK Biobank data are available upon application to the UK Biobank. Additional data that were used to generate the Figures in this study are available on request from the corresponding author (S.M.D.) or through dbGAP as listed above.

https://github.com/Bellomot/Athero_NGWAMA_Multitrait_GWAS

## ACKNOWLEDGEMENTS

We want to thank all of the patients who participated in the GWAS studies.

## SOURCES OF FUNDING

This research is based on data from the Million Veteran Program, Office of Research and Development, Veterans Health Administration, and was supported by awardno. MVP000. This publication does not represent the views of the Department of Veteran Affairs or the United States Government. This research was also supported by funding from: the Department of Veterans Affairs awards nos. I01-BX03340 (K.C. and P.W.F.W.), I01-BX003362 (P.S.T. and K.M.C) and IK2-CX001780 (S.M.D.), the National Institutes of Health (DK101478 and DK126194 to B.F.V.), the American Heart Association (20PRE35120109 to W.P.B.).

## DISCLOSURES

S.M.D. receives research support to his institution from CytoVAS and RenalytixAI. S.K. is a founder of Maze Therapeutics, Verve Therapeutics and San Therapeutics. He holds equity in Catabasis and San Therapeutics. He is a member of the scientific advisory boards for Regeneron Genetics Center and Corvidia Therapeutics; served as a consultant for Acceleron, Eli Lilly, Novartis, Merck, NovoNordisk, Novo Ventures, Ionis, Alnylam, Aegerion, Huag Partners, Noble Insights, Leerink Partners, Bayer Healthcare, Illumina, Color Genomics, MedGenome, Quest and Medscape; and reports patents related to a method of identifying and treating a person having a predisposition to or afflicted with cardiometabolic disease (20180010185) and a genetics risk predictor (20190017119). C.J.O. is employed by Novartis Institutes of Biomedical Research. All other authors have no conflicts of interest to declare.

## Notes

### Clinical Trial

The analysis in this manuscript was performed on publicly available data from human subject studies.

### Author Declarations

This study was reviewed and determined to be exempt by the Veteran's Association Central IRB. All data used in this manuscript were publicly available.

## REFERENCES

1. Kobiyama K, Ley K. Atherosclerosis. Circ Res. 2018;123(10):1118–1120.

2. Virani SS, Alonso A, Benjamin EJ, et al. Heart Disease and Stroke Statistics-2020 Update: A Report From the American Heart Association. Circulation. 2020;141(9):e139–e596.

3. Lozano R, Naghavi M, Foreman K, et al. Global and regional mortality from 235 causes of death for 20 age groups in 1990 and 2010: a systematic analysis for the Global Burden of Disease Study 2010. Lancet. 2012;380(9859):2095–2128.

4. Lin JS, Olson CM, Johnson ES, Whitlock EP. The ankle-brachial index for peripheral artery disease screening and cardiovascular disease prediction among asymptomatic adults: a systematic evidence review for the U.S. Preventive Services Task Force. Ann Intern Med. 2013;159(5):333–341.

5. Sundaram V, Bloom C, Zakeri R, et al. Temporal trends in the incidence, treatment patterns, and outcomes of coronary artery disease and peripheral artery disease in the UK, 2006-2015. Eur Heart J. 2020;41(17):1636–1649.

6. Klarin D, Lynch J, Aragam K, et al. Genome-wide association study of peripheral artery disease in the Million Veteran Program. Nat Med. 2019;25(8):1274–1279.

7. Ozkaramanli Gur D, Guzel S, Akyuz A, Alpsoy S, Guler N. The role of novel cytokines in inflammation: Defining peripheral artery disease among patients with coronary artery disease. Vasc Med. 2018;23(5):428–436.

8. Criqui MH, Aboyans V. Epidemiology of peripheral artery disease. Circ Res. 2015;116(9):1509–1526.

9. Hussein AA, Uno K, Wolski K, et al. Peripheral arterial disease and progression of coronary atherosclerosis. J Am Coll Cardiol. 2011;57(10):1220–1225.

10. van der Harst P, Verweij N. Identification of 64 Novel Genetic Loci Provides an Expanded View on the Genetic Architecture of Coronary Artery Disease. Circ Res. 2018;122(3):433–443.

11. Khera AV, Kathiresan S. Genetics of coronary artery disease: discovery, biology and clinical translation. Nat Rev Genet. 2017;18(6):331–344.

12. Koyama S, Ito K, Terao C, et al. Population-specific and trans-ancestry genome-wide analyses identify distinct and shared genetic risk loci for coronary artery disease. Nat Genet. 2020;52(11):1169–1177.

13. Matsukura M, Ozaki K, Takahashi A, et al. Genome-Wide Association Study of Peripheral Arterial Disease in a Japanese Population. PLoS One. 2015;10(10):e0139262.

14. Purcell S, Neale B, Todd-Brown K, et al. PLINK: a tool set for whole-genome association and population-based linkage analyses. Am J Hum Genet. 2007;81(3):559–575.

15. Baselmans BML, Jansen R, Ip HF, et al. Multivariate genome-wide analyses of the well-being spectrum. Nat Genet. 2019;51(3):445–451.

16. Zhao W, Rasheed A, Tikkanen E, et al. Identification of new susceptibility loci for type 2 diabetes and shared etiological pathways with coronary heart disease. Nat Genet. 2017;49(10):1450–1457.

17. Siewert KM, Voight BF. Bivariate Genome-Wide Association Scan Identifies 6 Novel Loci Associated With Lipid Levels and Coronary Artery Disease. Circ Genom Precis Med. 2018;11(12):e002239.

18. Holmes MV, Asselbergs FW, Palmer TM, et al. Mendelian randomization of blood lipids for coronary heart disease. Eur Heart J. 2015;36(9):539–550.

19. Riaz H, Khan MS, Siddiqi TJ, et al. Association Between Obesity and Cardiovascular Outcomes: A Systematic Review and Meta-analysis of Mendelian Randomization Studies. JAMA Netw Open. 2018;1(7):e183788.

20. Larsson SC, Mason AM, Back M, et al. Genetic predisposition to smoking in relation to 14 cardiovascular diseases. Eur Heart J. 2020;41(35):3304–3310.

21. Yengo L, Sidorenko J, Kemper KE, et al. Meta-analysis of genome-wide association studies for height and body mass index in approximately 700000 individuals of European ancestry. Hum Mol Genet. 2018;27(20):3641–3649.

22. Vujkovic M, Keaton JM, Lynch JA, et al. Discovery of 318 new risk loci for type 2 diabetes and related vascular outcomes among 1.4 million participants in a multi-ancestry meta-analysis. Nat Genet. 2020;52(7):680–691.

23. Wootton RE, Richmond RC, Stuijfzand BG, et al. Evidence for causal effects of lifetime smoking on risk for depression and schizophrenia: a Mendelian randomisation study. Psychol Med. 2020;50(14):2435–2443.

24. Klarin D, Damrauer SM, Cho K, et al. Genetics of blood lipids among ∼300,000 multiethnic participants of the Million Veteran Program. Nat Genet. 2018;50(11):1514–1523.

25. Buniello A, MacArthur JAL, Cerezo M, et al. The NHGRI-EBI GWAS Catalog of published genome-wide association studies, targeted arrays and summary statistics 2019. Nucleic Acids Res. 2019;47(D1):D1005–D1012.

26. Giambartolomei C, Vukcevic D, Schadt EE, et al. Bayesian test for colocalisation between pairs of genetic association studies using summary statistics. PLoS Genet. 2014;10(5):e1004383.

27. Giambartolomei C, Zhenli Liu J, Zhang W, et al. A Bayesian framework for multiple trait colocalization from summary association statistics. Bioinformatics. 2018;34(15):2538–2545.

28. Pruim RJ, Welch RP, Sanna S, et al. LocusZoom: regional visualization of genome-wide association scan results. Bioinformatics. 2010;26(18):2336–2337.

29. Yang J, Lee SH, Goddard ME, Visscher PM. GCTA: a tool for genome-wide complex trait analysis. Am J Hum Genet. 2011;88(1):76–82.

30. Genomes Project C, Abecasis GR, Auton A, et al. An integrated map of genetic variation from 1,092 human genomes. Nature. 2012;491(7422):56–65.

31. K.A.B. Gawronski WB, Y. Park, E. Pashos, X. Wang, W. Yang, D. Rader, K. Musunuru, B. Voight, C. Brown. Evaluating the contribution of cell-type specific alternative splicing to variation in lipid levels. biorxiv. 2019.

32. Castel SE, Aguet F, Mohammadi P, Consortium GT, Ardlie KG, Lappalainen T. A vast resource of allelic expression data spanning human tissues. Genome Biol. 2020;21(1):234.

33. Liu M, Jiang Y, Wedow R, et al. Association studies of up to 1.2 million individuals yield new insights into the genetic etiology of tobacco and alcohol use. Nat Genet. 2019;51(2):237–244.

34. Klimentidis YC, Arora A, Newell M, et al. Phenotypic and Genetic Characterization of Lower LDL Cholesterol and Increased Type 2 Diabetes Risk in the UK Biobank. Diabetes. 2020;69(10):2194–2205.

35. Richardson TG, Sanderson E, Palmer TM, et al. Evaluating the relationship between circulating lipoprotein lipids and apolipoproteins with risk of coronary heart disease: A multivariable Mendelian randomisation analysis. PLoS Med. 2020;17(3):e1003062.

36. Ruotsalainen SE, Partanen JJ, Cichonska A, et al. An expanded analysis framework for multivariate GWAS connects inflammatory biomarkers to functional variants and disease. Eur J Hum Genet. 2021;29(2):309–324.

37. Iyengar SK, Sedor JR, Freedman BI, et al. Genome-Wide Association and Trans-ethnic Meta-Analysis for Advanced Diabetic Kidney Disease: Family Investigation of Nephropathy and Diabetes (FIND). PLoS Genet. 2015;11(8):e1005352.

38. Hackinger S, Prins B, Mamakou V, et al. Evidence for genetic contribution to the increased risk of type 2 diabetes in schizophrenia. Transl Psychiatry. 2018;8(1):252.

39. Nath AP, Ritchie SC, Grinberg NF, et al. Multivariate Genome-wide Association Analysis of a Cytokine Network Reveals Variants with Widespread Immune, Haematological, and Cardiometabolic Pleiotropy. Am J Hum Genet. 2019;105(6):1076–1090.

40. Folkersen L, Gustafsson S, Wang Q, et al. Genomic and drug target evaluation of 90 cardiovascular proteins in 30,931 individuals. Nat Metab. 2020;2(10):1135–1148.

41. Kettunen J, Demirkan A, Wurtz P, et al. Genome-wide study for circulating metabolites identifies 62 loci and reveals novel systemic effects of LPA. Nat Commun. 2016;7:11122.

42. Pashos EE, Park Y, Wang X, et al. Large, Diverse Population Cohorts of hiPSCs and Derived Hepatocyte-like Cells Reveal Functional Genetic Variation at Blood Lipid-Associated Loci. Cell Stem Cell. 2017;20(4):558–570 e510.

43. Pulit SL, Stoneman C, Morris AP, et al. Meta-analysis of genome-wide association studies for body fat distribution in 694 649 individuals of European ancestry. Hum Mol Genet. 2019;28(1):166–174.

44. Zhu Z, Guo Y, Shi H, et al. Shared genetic and experimental links between obesity-related traits and asthma subtypes in UK Biobank. J Allergy Clin Immunol. 2020;145(2):537–549.

45. Turcot V, Lu Y, Highland HM, et al. Protein-altering variants associated with body mass index implicate pathways that control energy intake and expenditure in obesity. Nat Genet. 2018;50(1):26–41.

46. Anflous K, Armstrong DD, Craigen WJ. Altered mitochondrial sensitivity for ADP and maintenance of creatine-stimulated respiration in oxidative striated muscles from VDAC1-deficient mice. J Biol Chem. 2001;276(3):1954–1960.

47. Subedi KP, Kim JC, Kang M, Son MJ, Kim YS, Woo SH. Voltage-dependent anion channel 2 modulates resting Ca(2)+ sparks, but not action potential-induced Ca(2)+ signaling in cardiac myocytes. Cell Calcium. 2011;49(2):136–143.

48. Alvira CM, Umesh A, Husted C, et al. Voltage-dependent anion channel-2 interaction with nitric oxide synthase enhances pulmonary artery endothelial cell nitric oxide production. Am J Respir Cell Mol Biol. 2012;47(5):669–678.

49. Gagarin D, Yang Z, Butler J, et al. Genomic profiling of acquired resistance to apoptosis in cells derived from human atherosclerotic lesions: potential role of STATs, cyclinD1, BAD, and Bcl-XL. J Mol Cell Cardiol. 2005;39(3):453–465.

50. Bjorkerud S, Bjorkerud B. Apoptosis is abundant in human atherosclerotic lesions, especially in inflammatory cells (macrophages and T cells), and may contribute to the accumulation of gruel and plaque instability. Am J Pathol. 1996;149(2):367–380.

51. Cheng EH, Sheiko TV, Fisher JK, Craigen WJ, Korsmeyer SJ. VDAC2 inhibits BAK activation and mitochondrial apoptosis. Science. 2003;301(5632):513–517.

52. Siragusa M, Sessa WC. Telmisartan exerts pleiotropic effects in endothelial cells and promotes endothelial cell quiescence and survival. Arterioscler Thromb Vasc Biol. 2013;33(8):1852–1860.

53. Ye F, Wang J, Meng W, Qian J, Jin M. Proteomic investigation of effects of hydroxysafflor yellow A in oxidized low-density lipoprotein-induced endothelial injury. Sci Rep. 2017;7(1):17981.

54. Wilting F, Kopp R, Gurnev PA, et al. The antiarrhythmic compound efsevin directly modulates voltage-dependent anion channel 2 by binding to its inner wall and enhancing mitochondrial Ca(2+) uptake. Br J Pharmacol. 2020;177(13):2947–2958.

55. Dixon SJ, Lemberg KM, Lamprecht MR, et al. Ferroptosis: an iron-dependent form of nonapoptotic cell death. Cell. 2012;149(5):1060–1072.

56. Yagoda N, von Rechenberg M, Zaganjor E, et al. RAS-RAF-MEK-dependent oxidative cell death involving voltage-dependent anion channels. Nature. 2007;447(7146):864–868.

57. Kiefer MC, Tucker JE, Joh R, Landsberg KE, Saltman D, Barr PJ. Identification of a second human subtilisin-like protease gene in the fes/fps region of chromosome 15. DNA Cell Biol. 1991;10(10):757–769.

58. Perisic L, Hedin E, Razuvaev A, et al. Profiling of atherosclerotic lesions by gene and tissue microarrays reveals PCSK6 as a novel protease in unstable carotid atherosclerosis. Arterioscler Thromb Vasc Biol. 2013;33(10):2432–2443.

59. Rykaczewska U, Suur BE, Rohl S, et al. PCSK6 Is a Key Protease in the Control of Smooth Muscle Cell Function in Vascular Remodeling. Circ Res. 2020;126(5):571–585.

60. Jin W, Fuki IV, Seidah NG, Benjannet S, Glick JM, Rader DJ. Proprotein convertases [corrected] are responsible for proteolysis and inactivation of endothelial lipase. J Biol Chem. 2005;280(44):36551–36559.

61. Choi S, Korstanje R. Proprotein convertases in high-density lipoprotein metabolism. Biomark Res. 2013;1(1):27.

62. Uniprot. UniProtKB - P29122 (PCSK6_HUMAN). https://www.uniprot.org/uniprot/P29122#P29122-1.

63. Ensembl. Transcript: PCSK6-215 ENST00000611967.4. https://useast.ensembl.org/Homo_sapiens/Transcript/ProteinSummary?db=core;g=ENSG00000140479;r=15:101297142-101525202;t=ENST00000611967.

64. Mori K, Kii S, Tsuji A, et al. A novel human PACE4 isoform, PACE4E is an active processing protease containing a hydrophobic cluster at the carboxy terminus. J Biochem. 1997;121(5):941–948.

65. Gauster M, Hrzenjak A, Schick K, Frank S. Endothelial lipase is inactivated upon cleavage by the members of the proprotein convertase family. J Lipid Res. 2005;46(5):977–987.

66. Jin W, Wang X, Millar JS, et al. Hepatic proprotein convertases modulate HDL metabolism. Cell Metab. 2007;6(2):129–136.

67. Byun S, Tortorella MD, Malfait AM, Fok K, Frank EH, Grodzinsky AJ. Transport and equilibrium uptake of a peptide inhibitor of PACE4 into articular cartilage is dominated by electrostatic interactions. Arch Biochem Biophys. 2010;499(1-2):32–39.

68. Komiyama T, Coppola JM, Larsen MJ, et al. Inhibition of furin/proprotein convertase-catalyzed surface and intracellular processing by small molecules. J Biol Chem. 2009;284(23):15729–15738.

