## Supplemental Figures 1-6 for "Multi-trait GWAS of atherosclerosis detects novel pleiotropic loci": Supp_Figures.pptx

### Slide 1
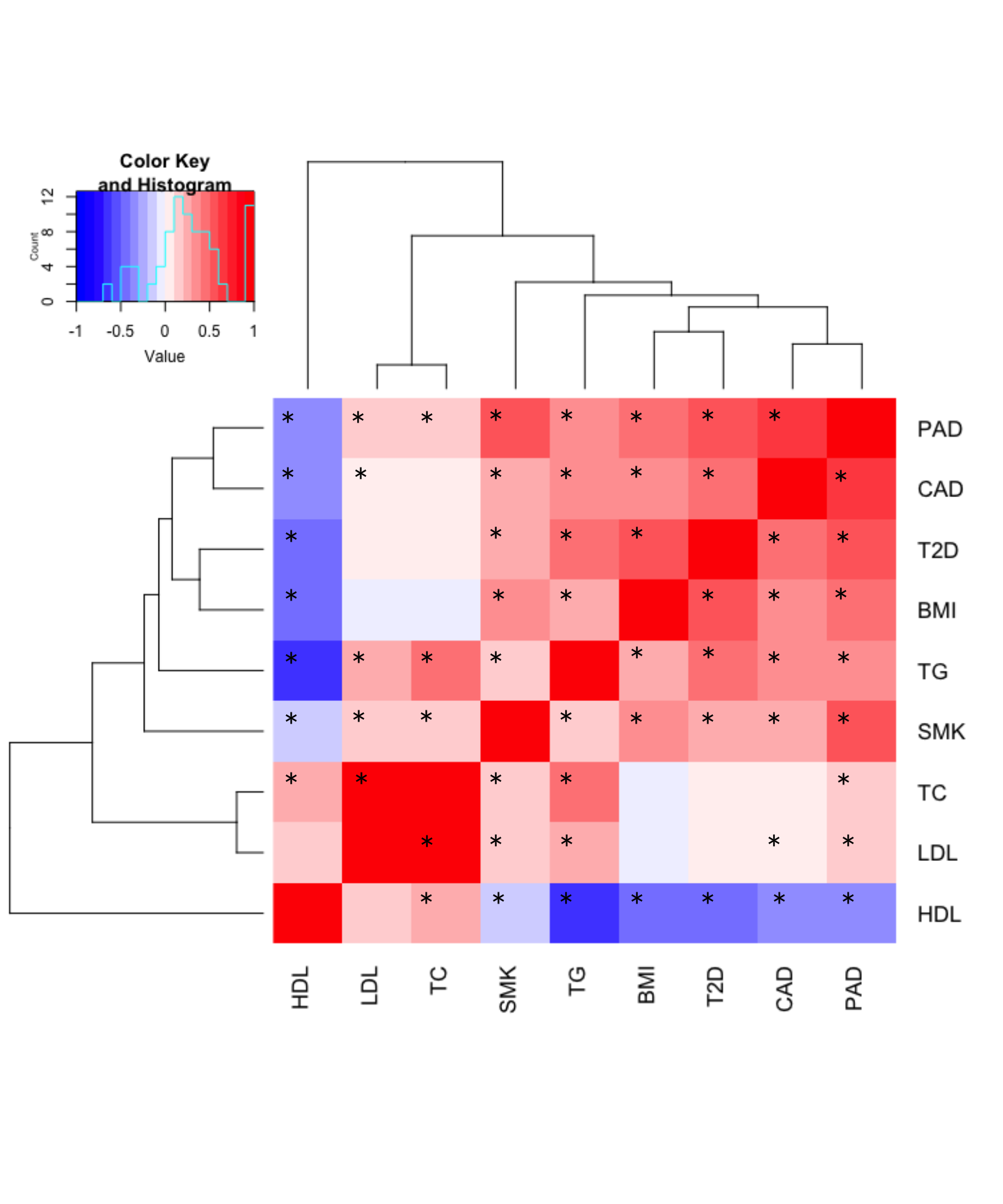

*
*
*
*
*
*
*
*
*
*
*
*
*
*
*
*
*
*
*
*
*
*
*
*
*
*
*
*
*
*
*
*
*
*
*
*
*
*
*
*
*
*
*
*
*
*
*
*
*
*
*
*
*
*
*
*
*
*
*
*

### Slide 2
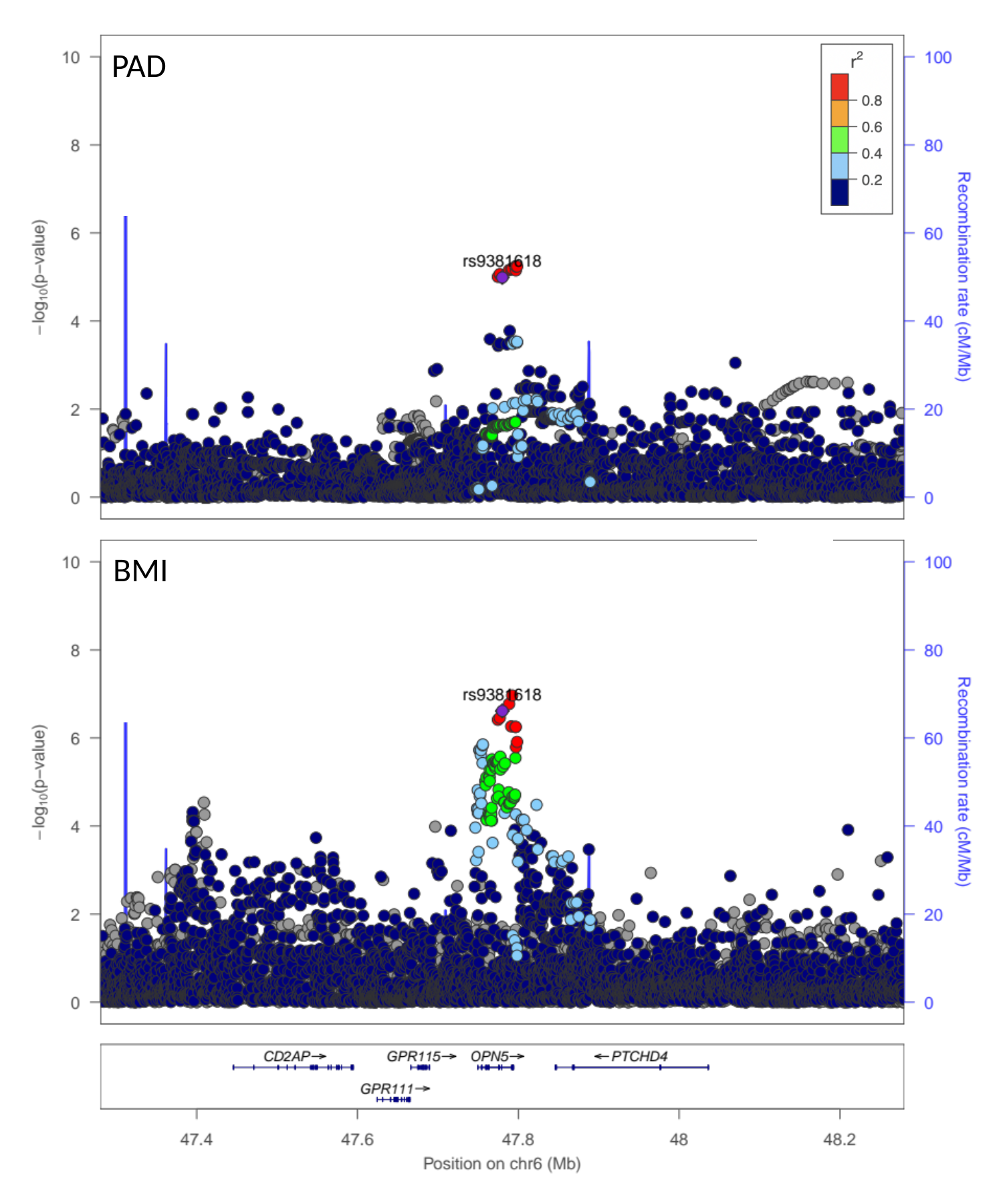

PAD
BMI

### Slide 3
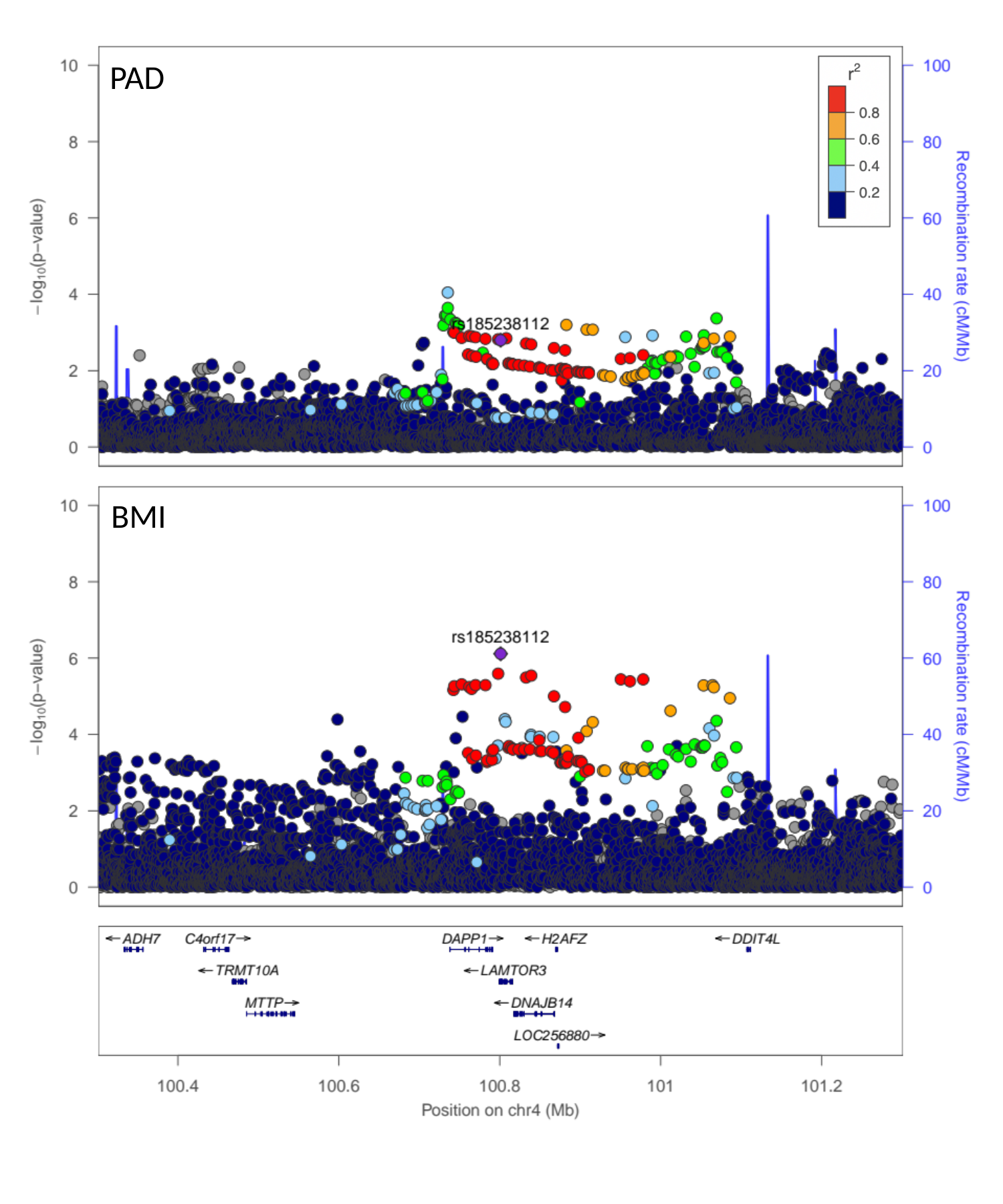

PAD
BMI

### Slide 4
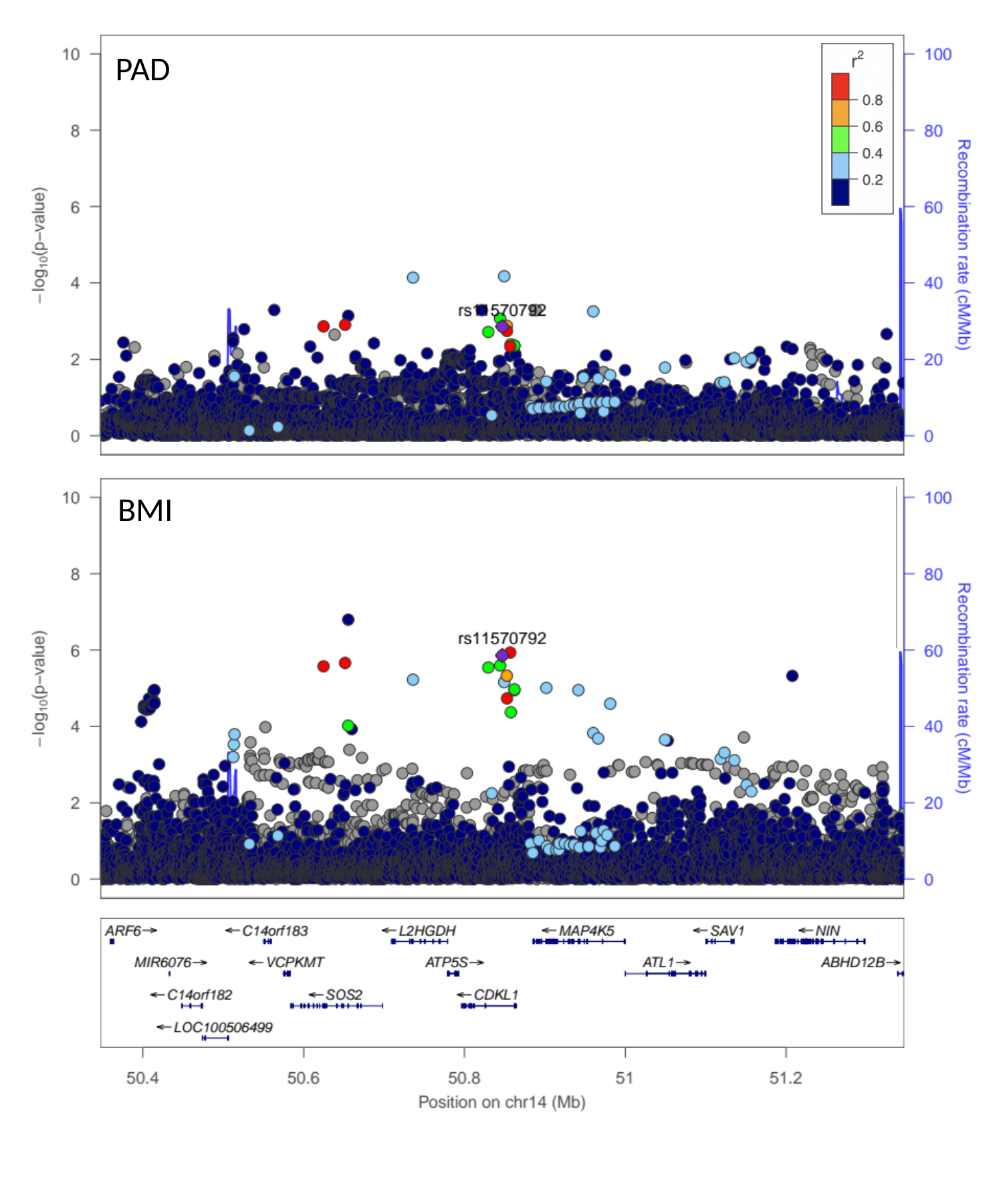

PAD
BMI

### Slide 5
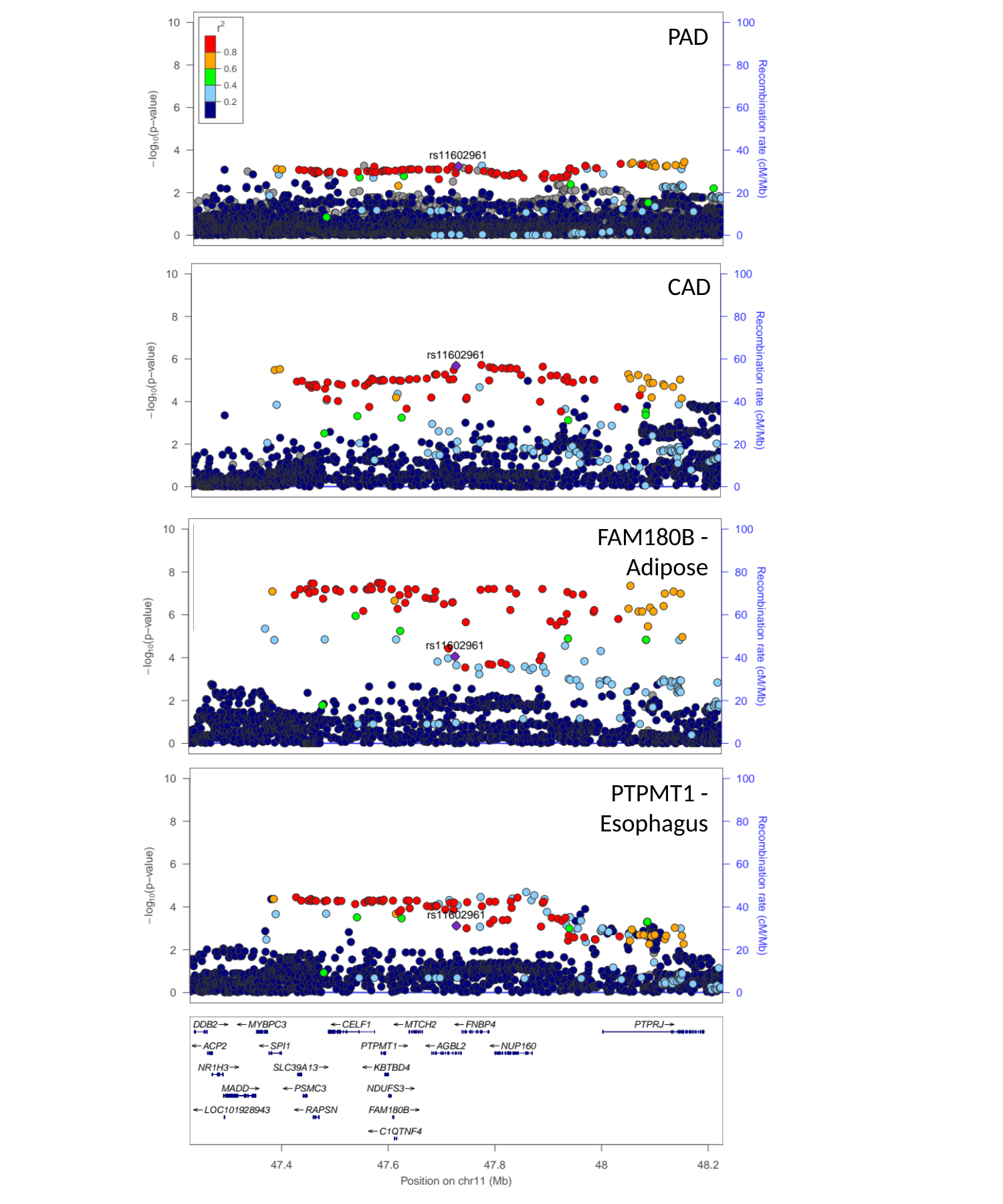

PAD
CAD
FAM180B -
Adipose
PTPMT1 -
Esophagus

### Slide 6
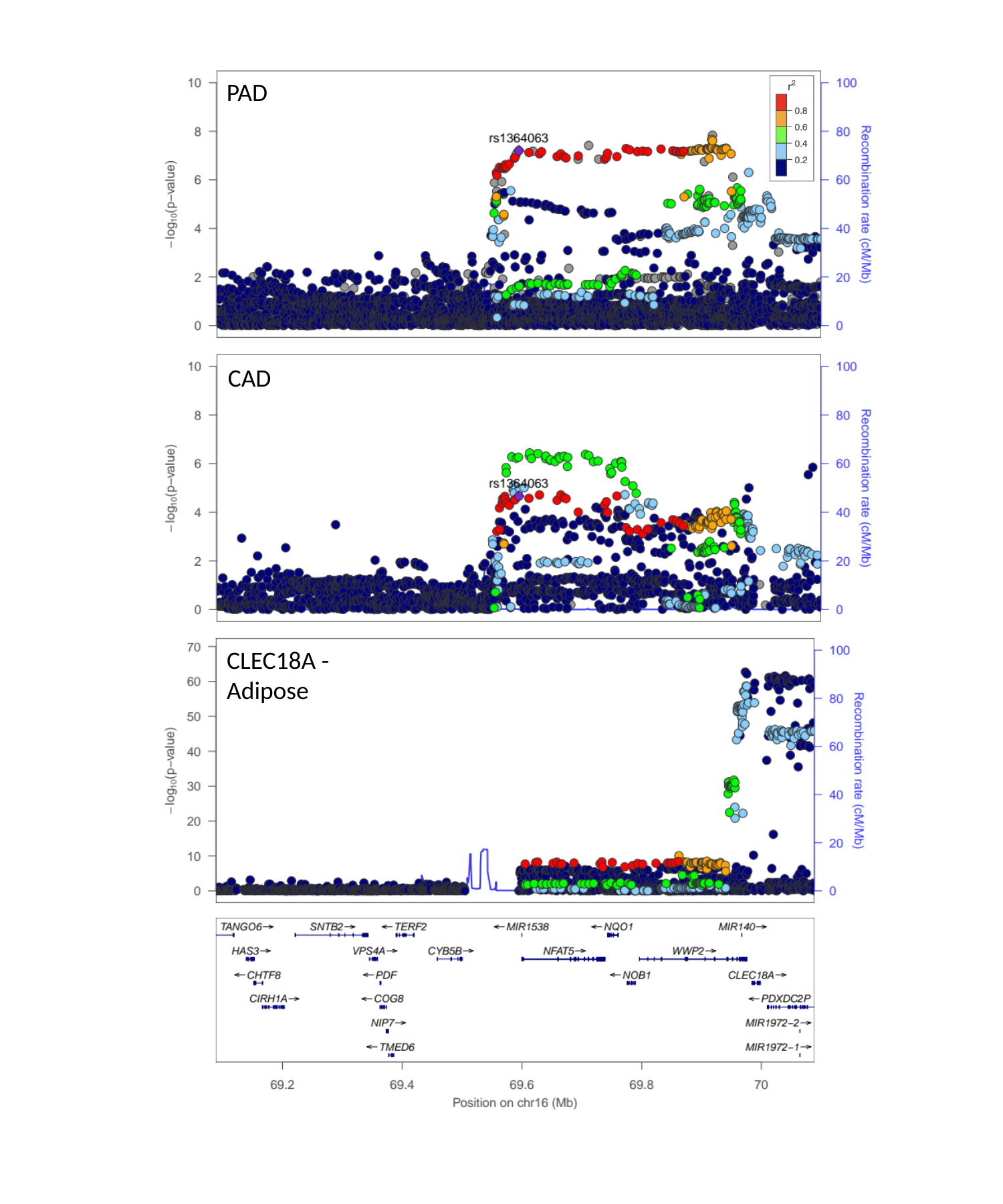

PAD
CAD
CLEC18A -
Adipose

### Slide 7
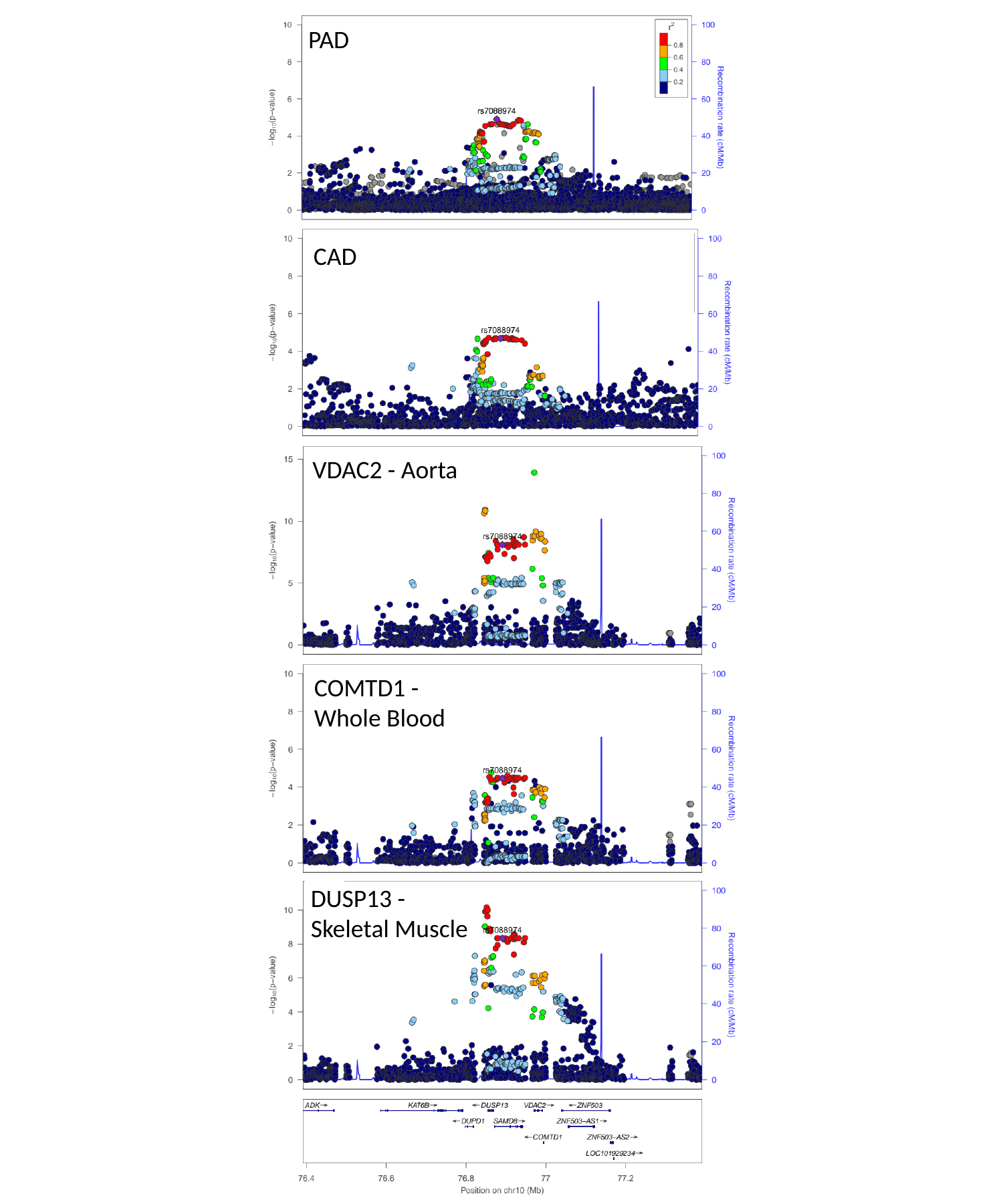

PAD
CAD
VDAC2 - Aorta
COMTD1 -
Whole Blood
DUSP13 -
Skeletal Muscle

### Slide 8
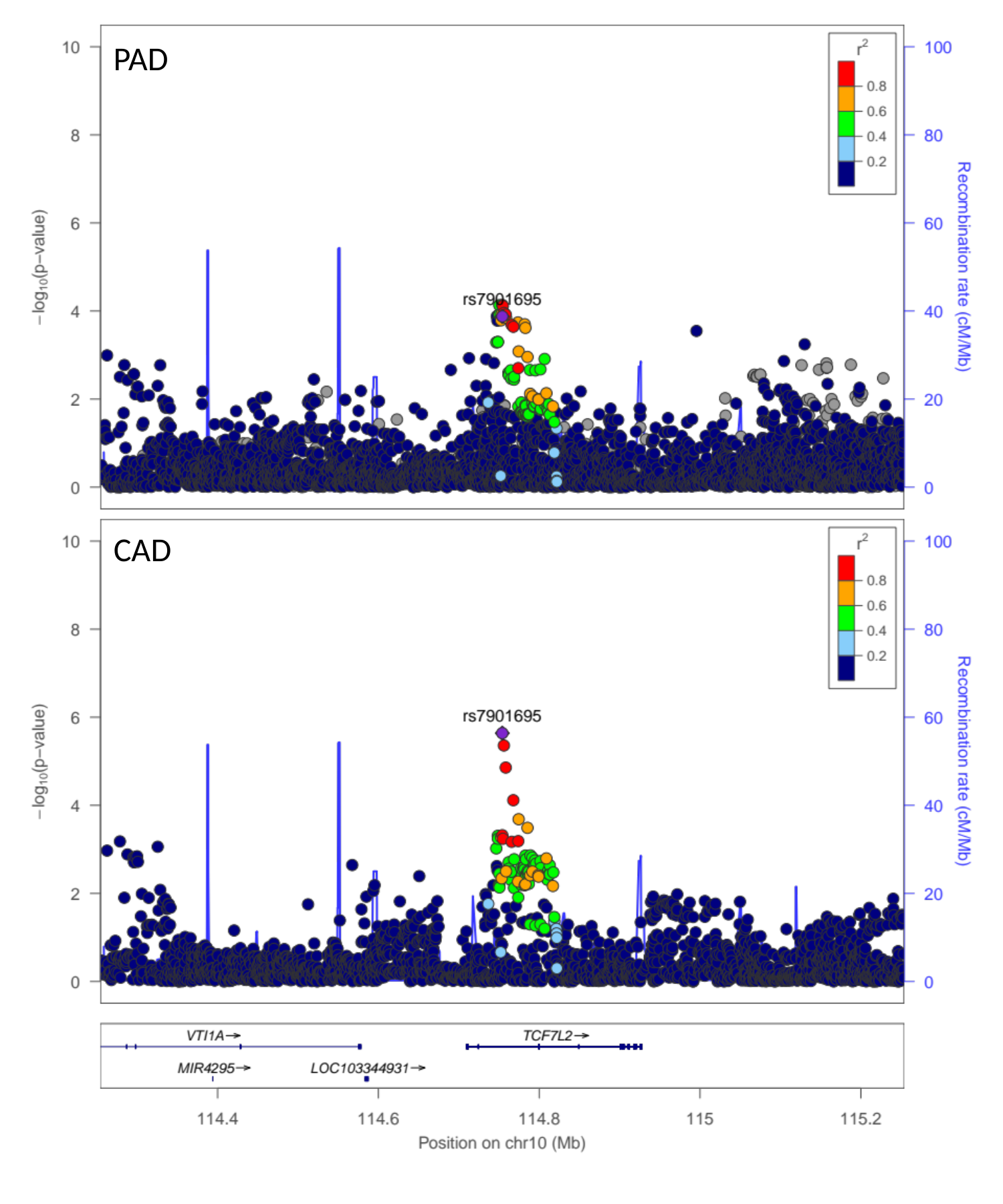

PAD
CAD

### Slide 9
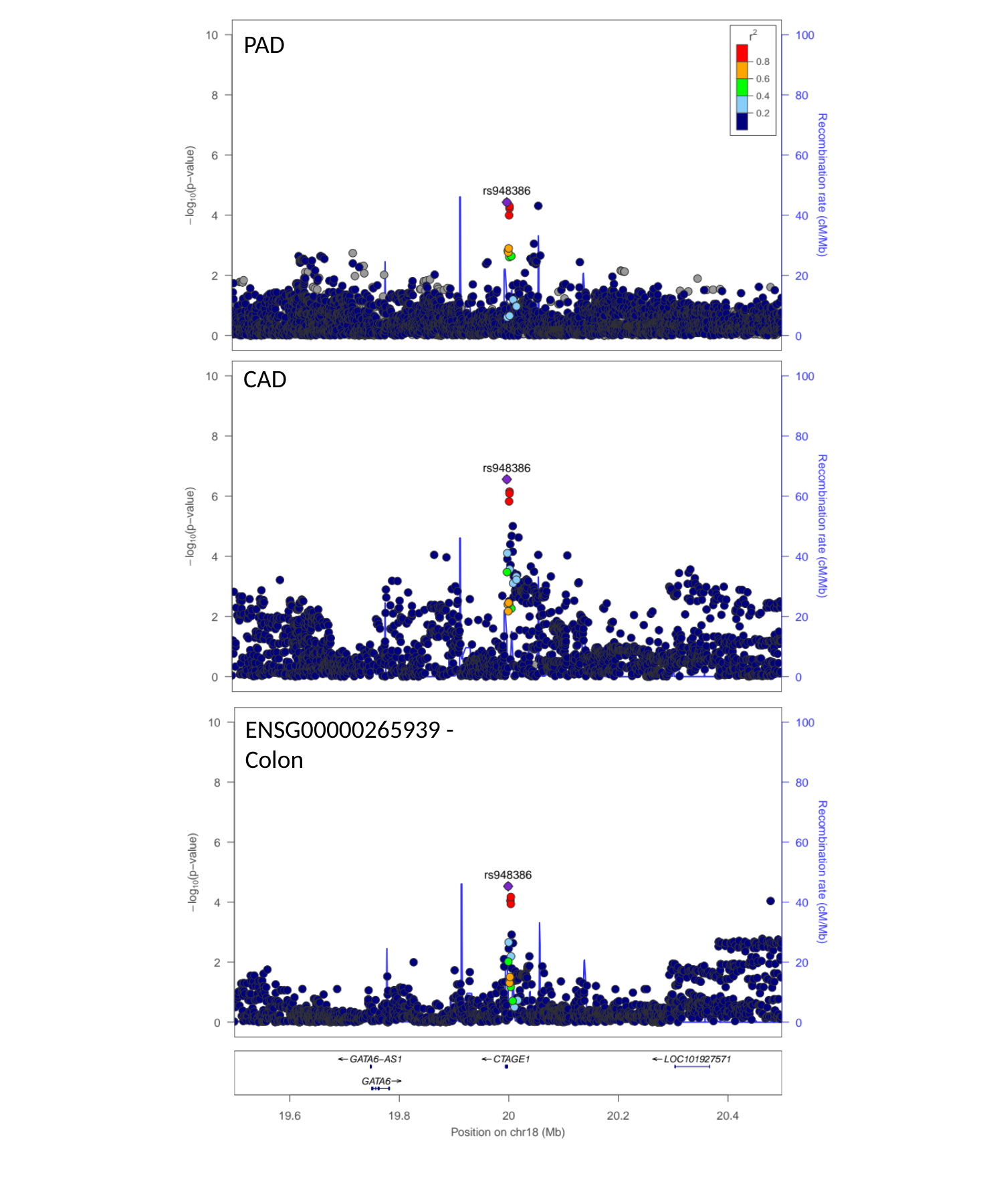

PAD
CAD
ENSG00000265939 -
Colon

### Slide 10
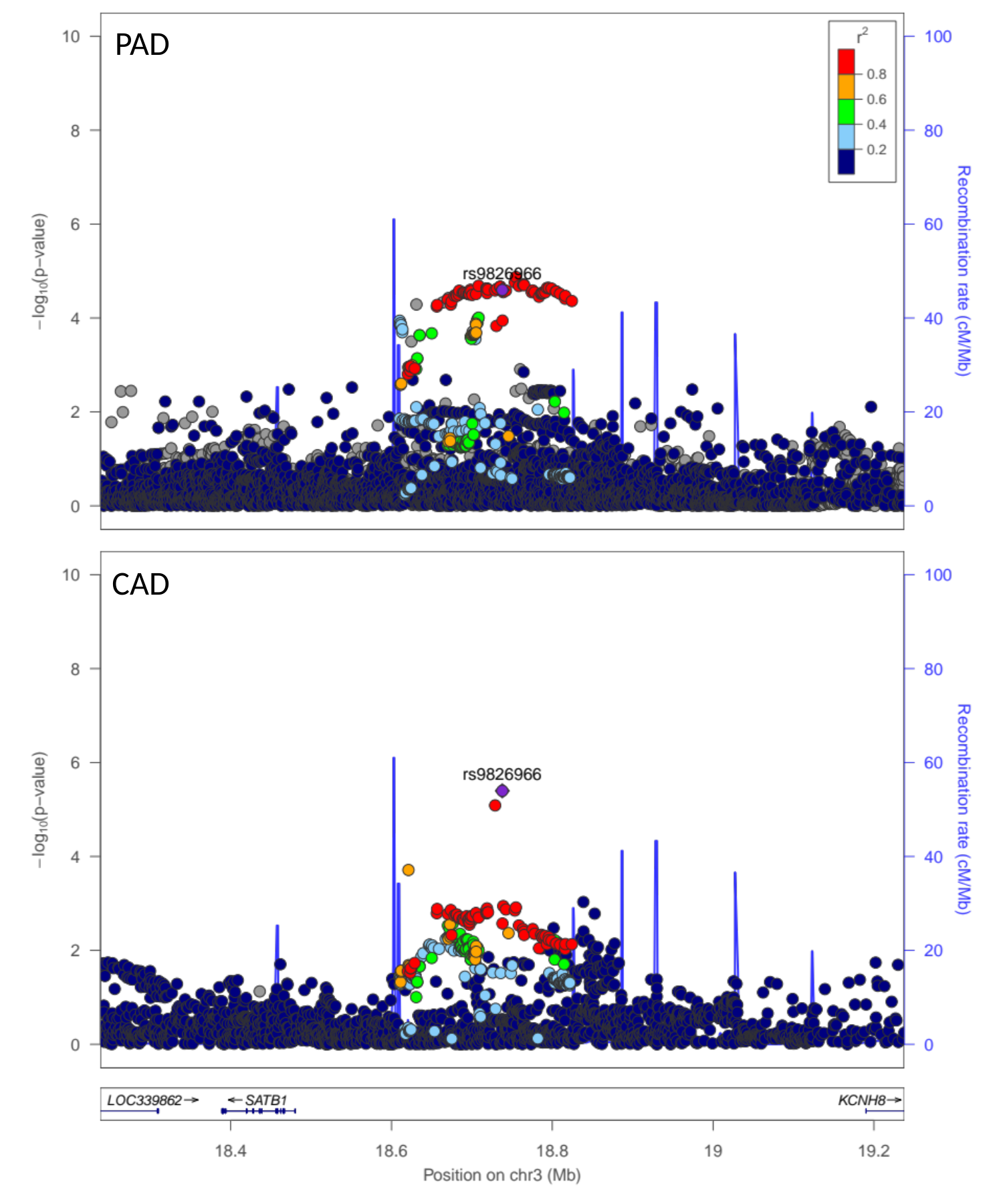

PAD
CAD

### Slide 11
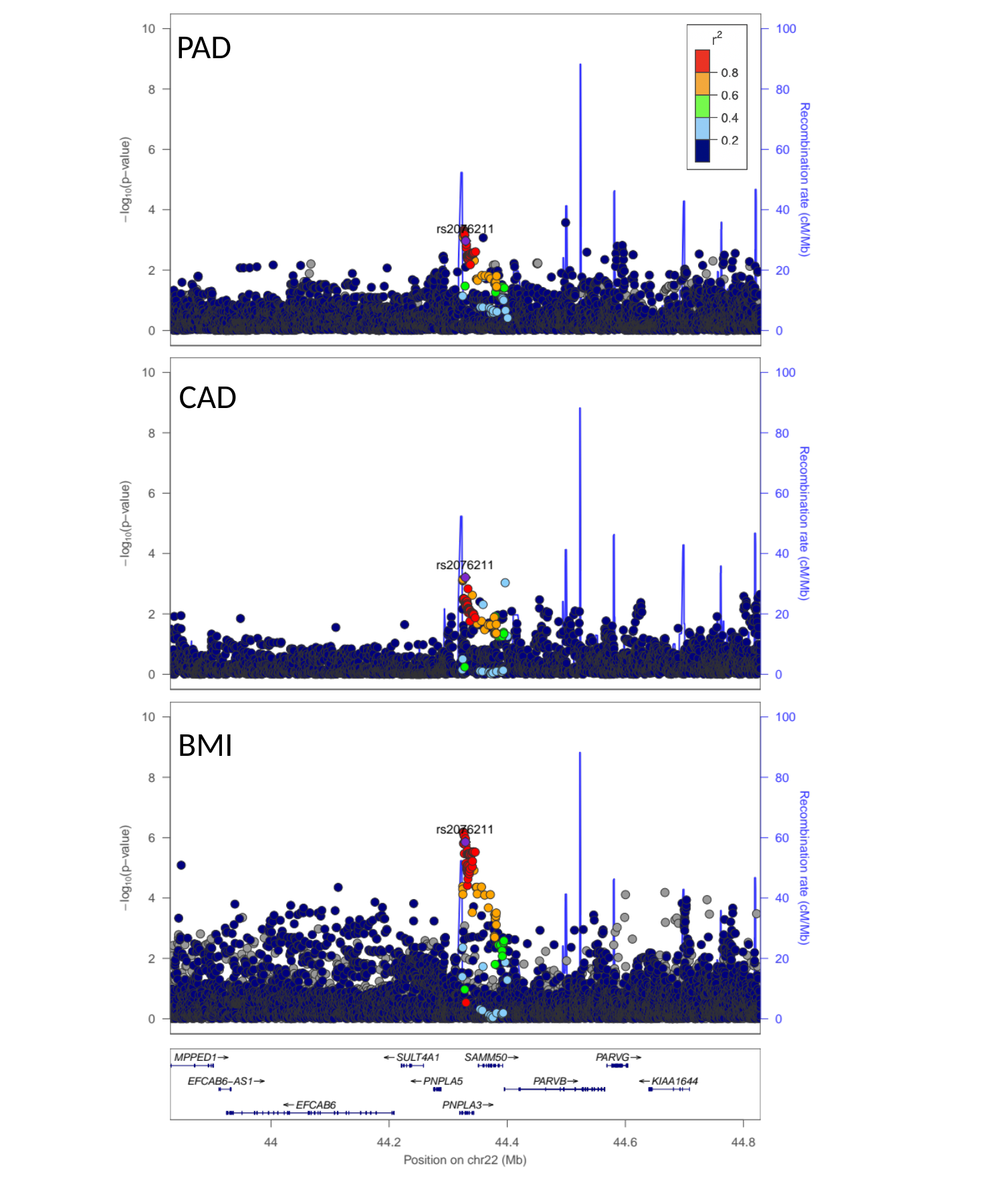

PAD
CAD
BMI

### Slide 12
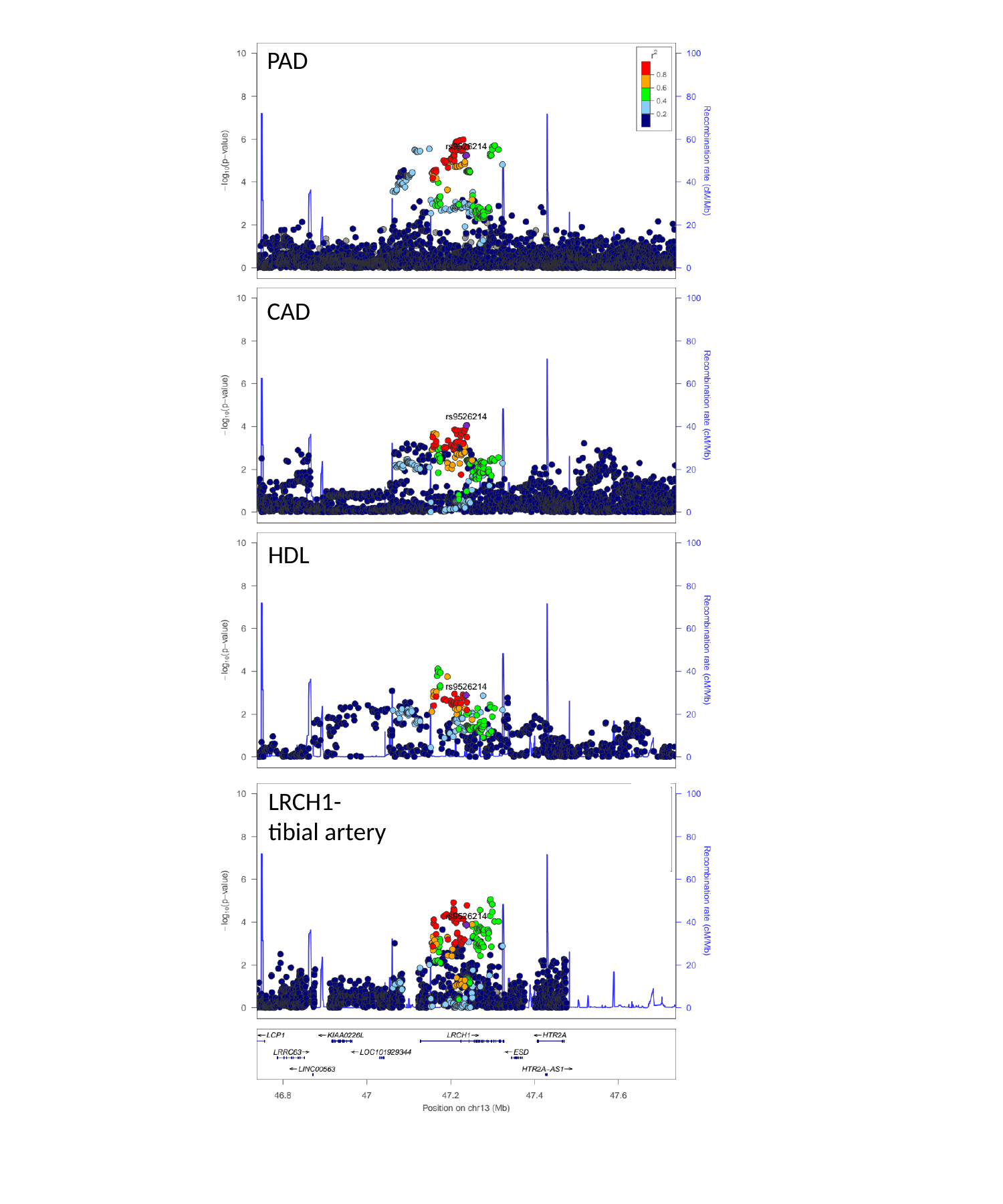

PAD
CAD
HDL
LRCH1-
tibial artery

### Slide 13
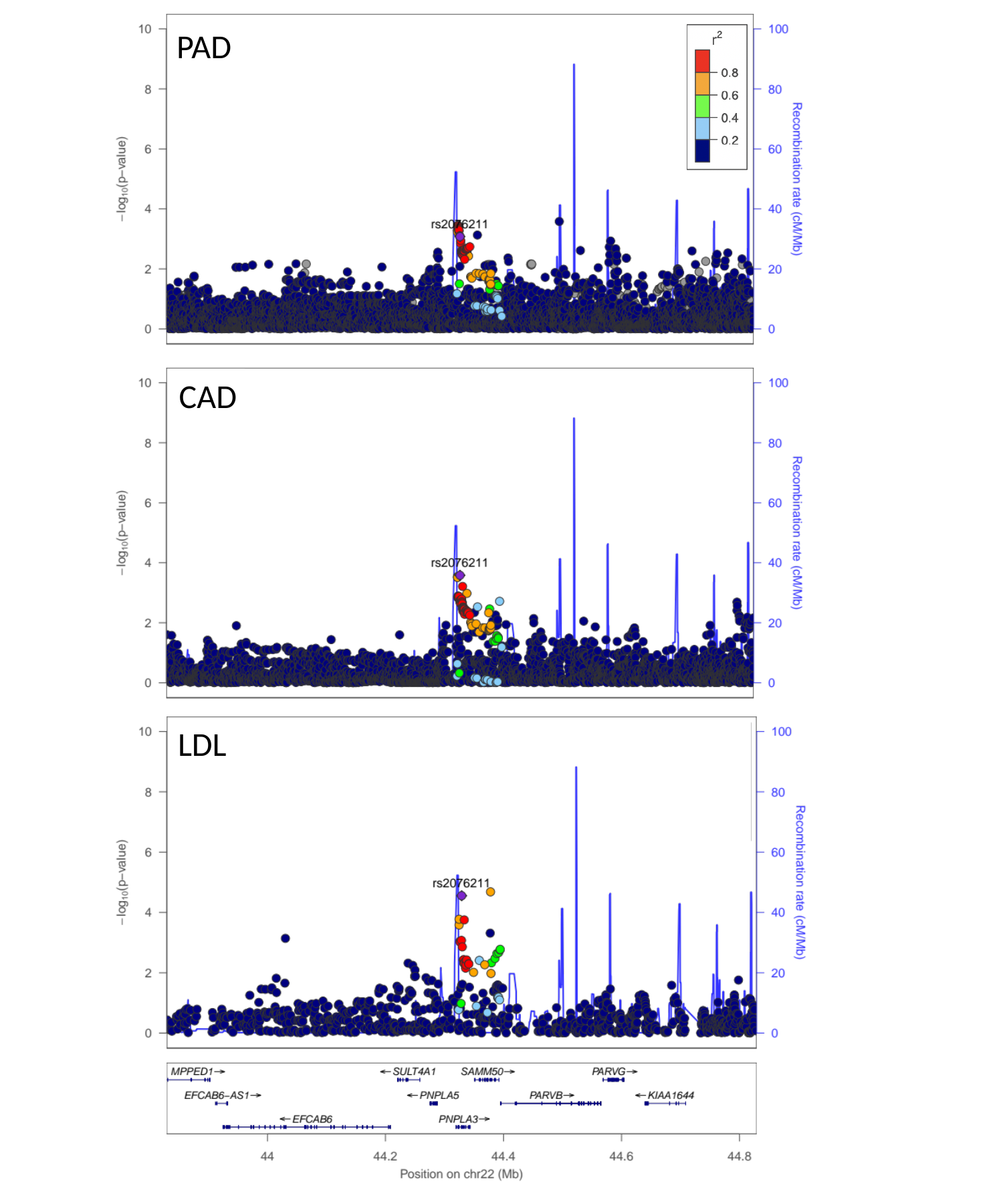

PAD
CAD
LDL

### Slide 14
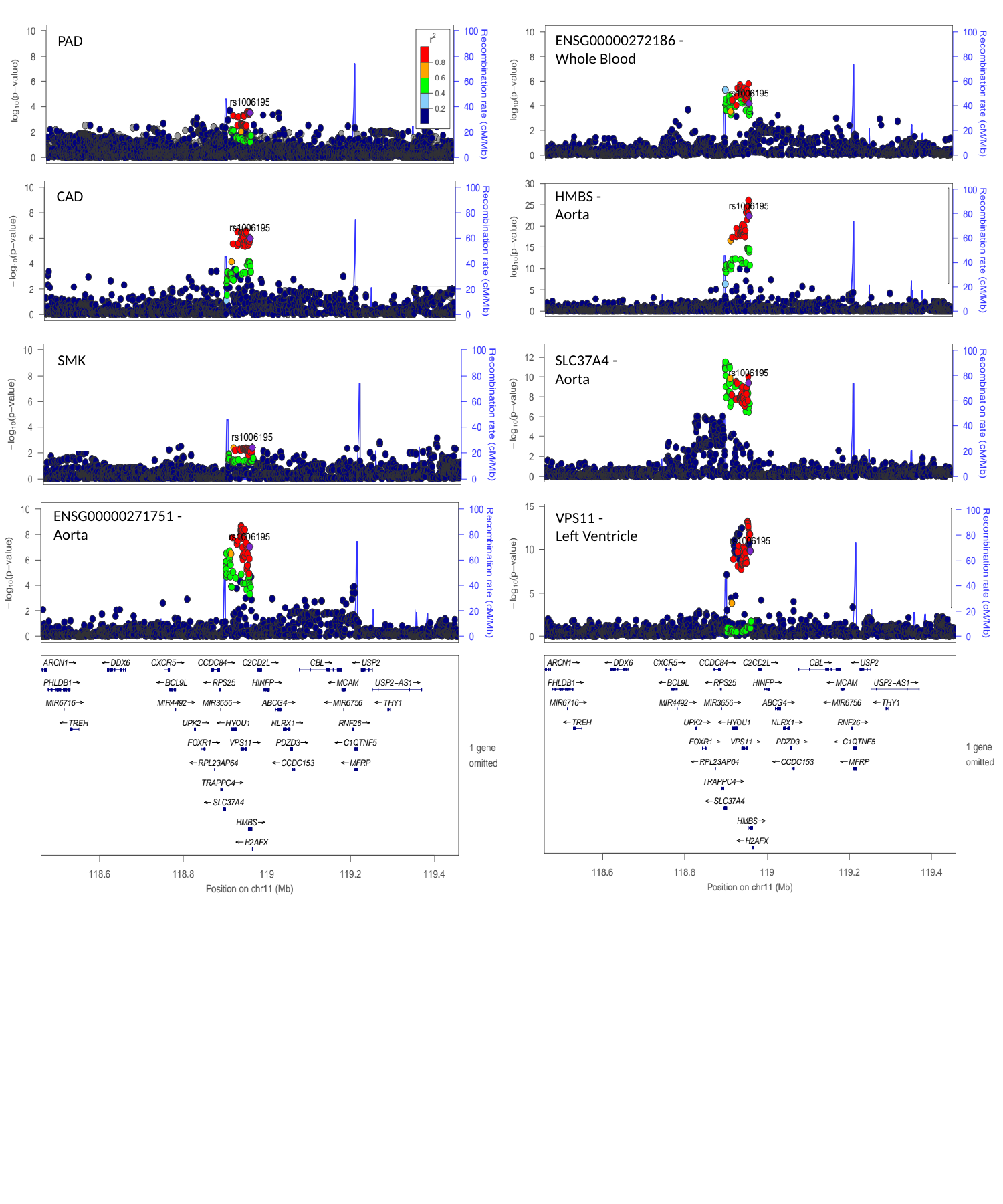

ENSG00000272186 -
Whole Blood
PAD
CAD
HMBS -
Aorta
SMK
SLC37A4 -
Aorta
ENSG00000271751 -
Aorta
VPS11 -
Left Ventricle

### Slide 15
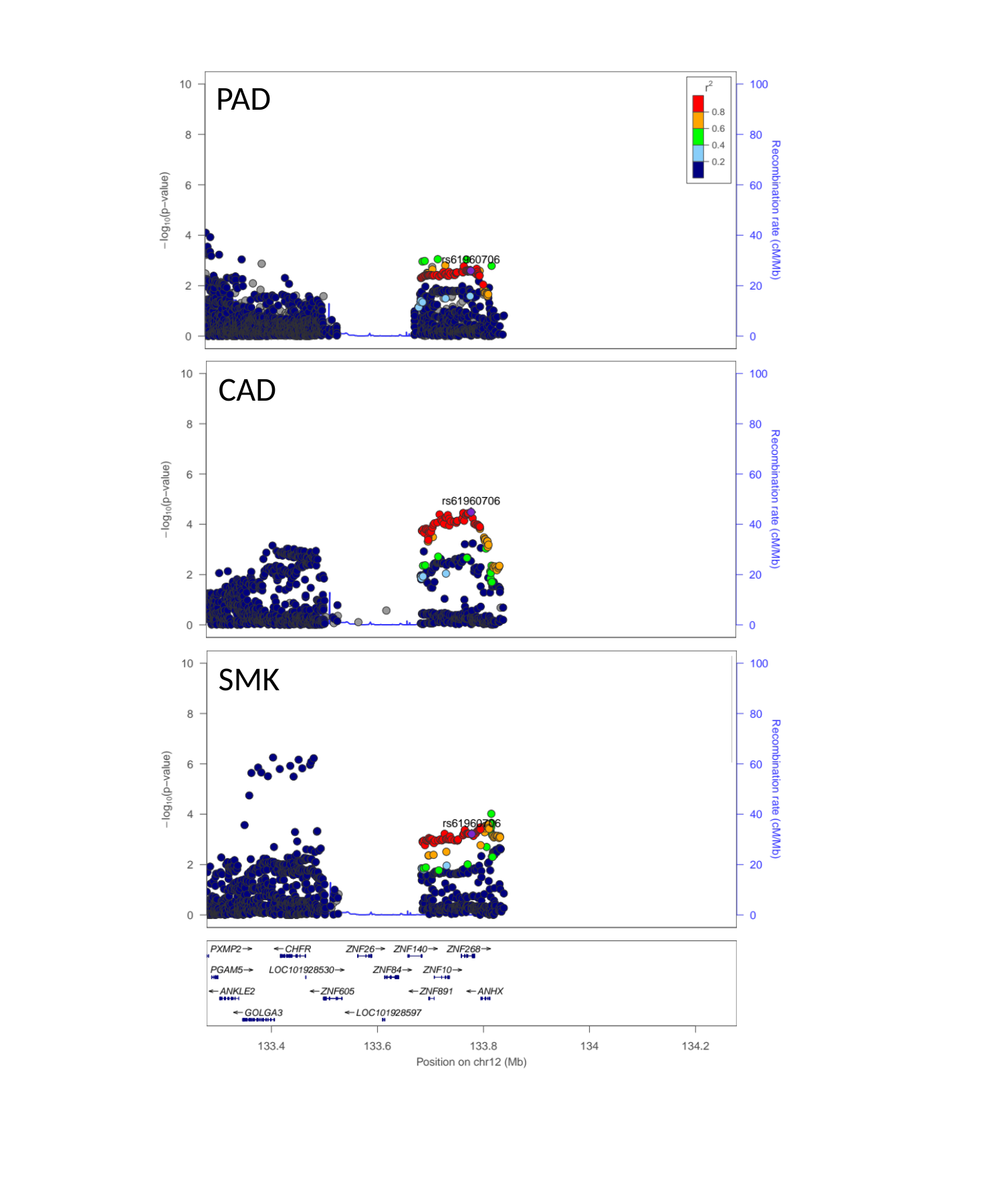

PAD
CAD
SMK

### Slide 16
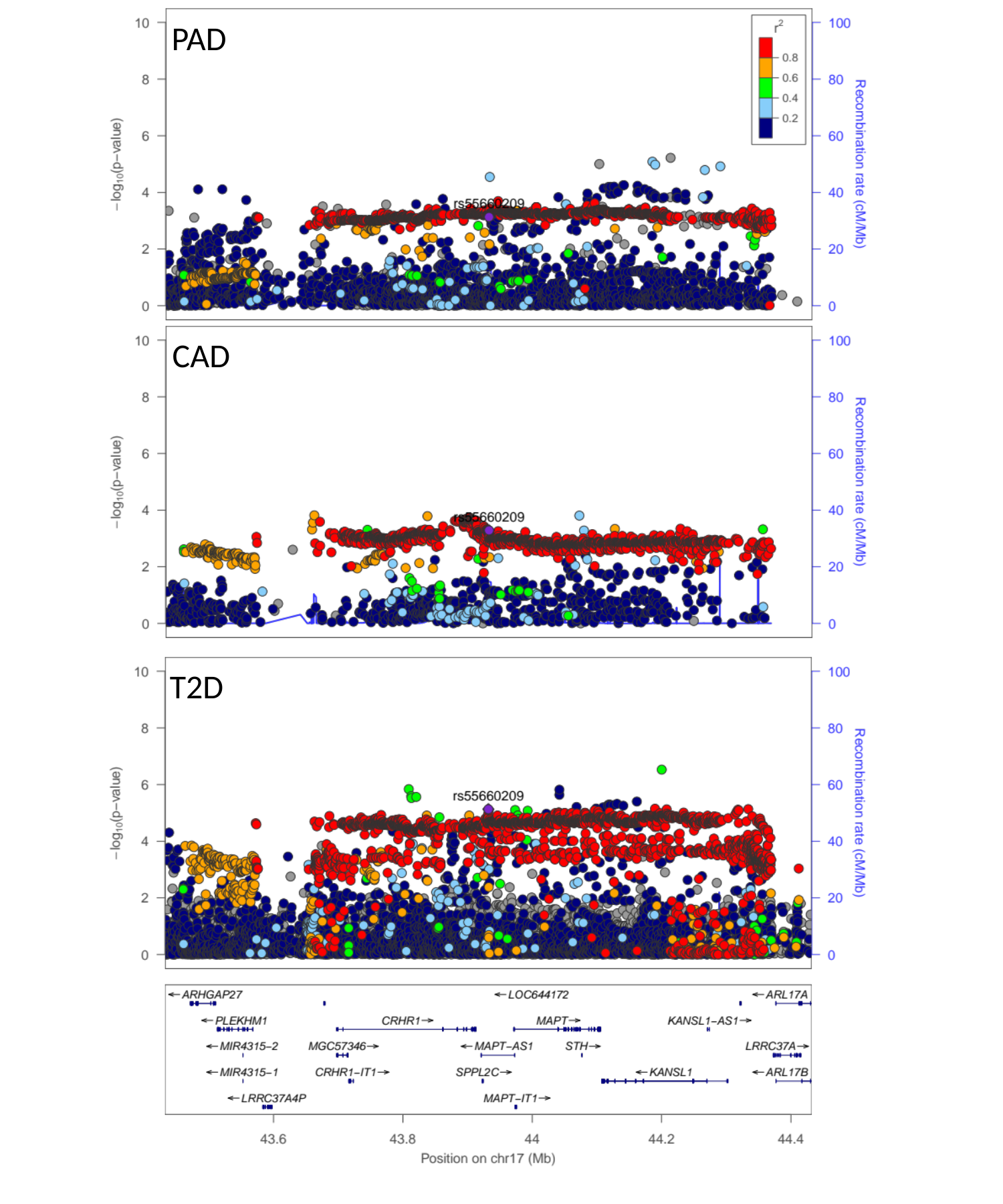

PAD
CAD
T2D

### Slide 17
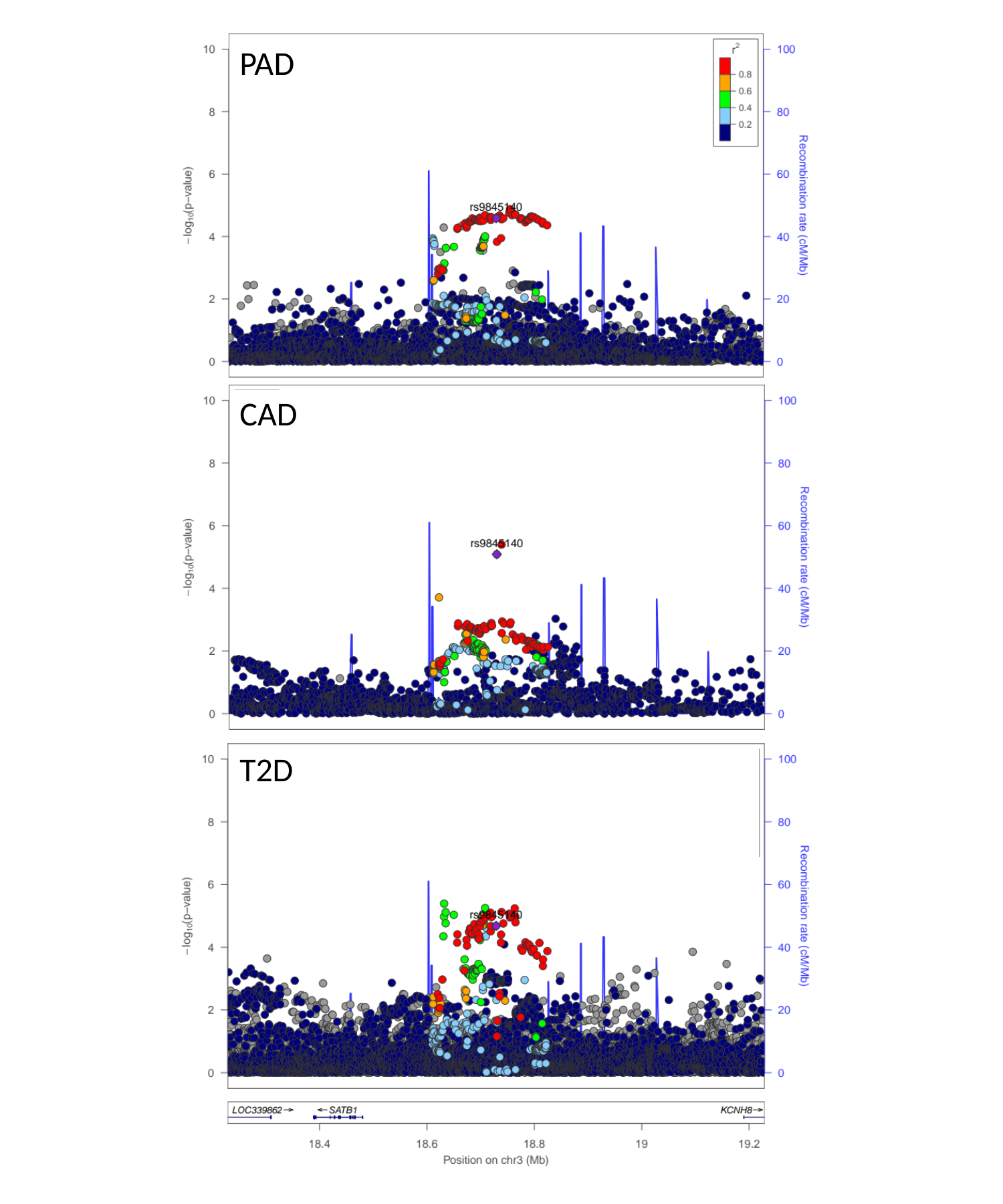

PAD
CAD
T2D

### Slide 18
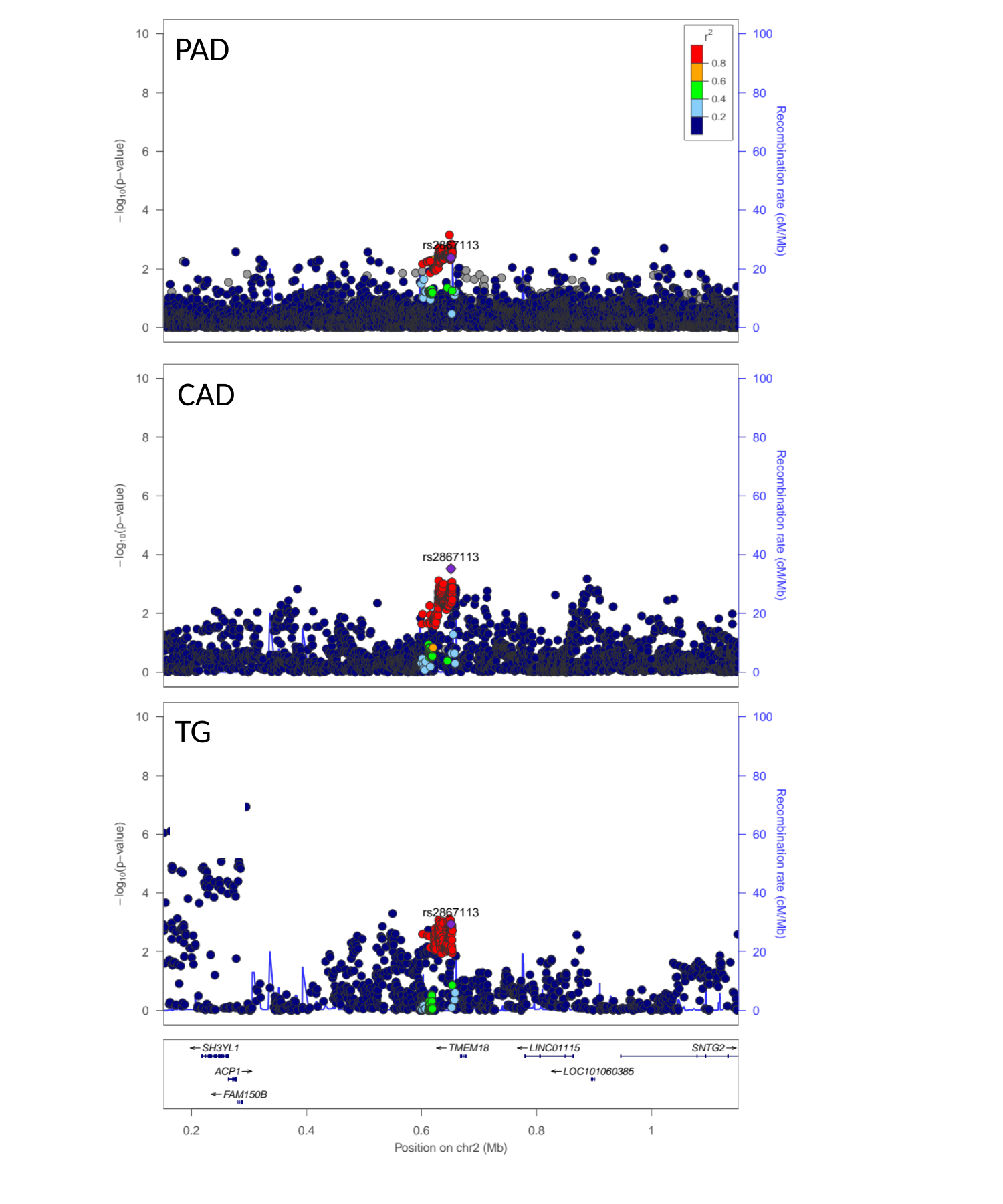

PAD
CAD
TG

### Slide 19
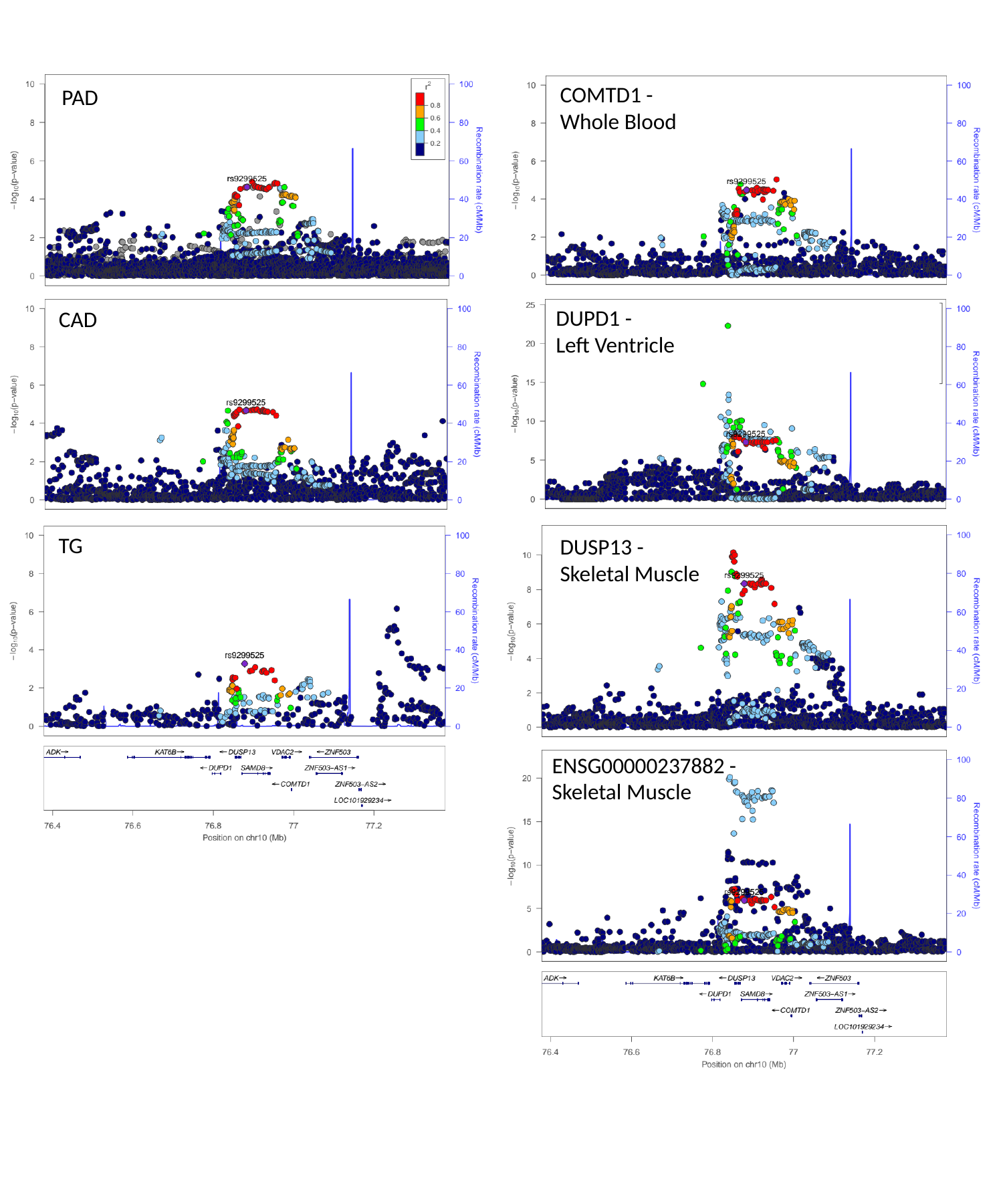

COMTD1 -
Whole Blood
PAD
DUPD1 -
Left Ventricle
CAD
TG
DUSP13 -
Skeletal Muscle
ENSG00000237882 -
Skeletal Muscle

### Slide 20
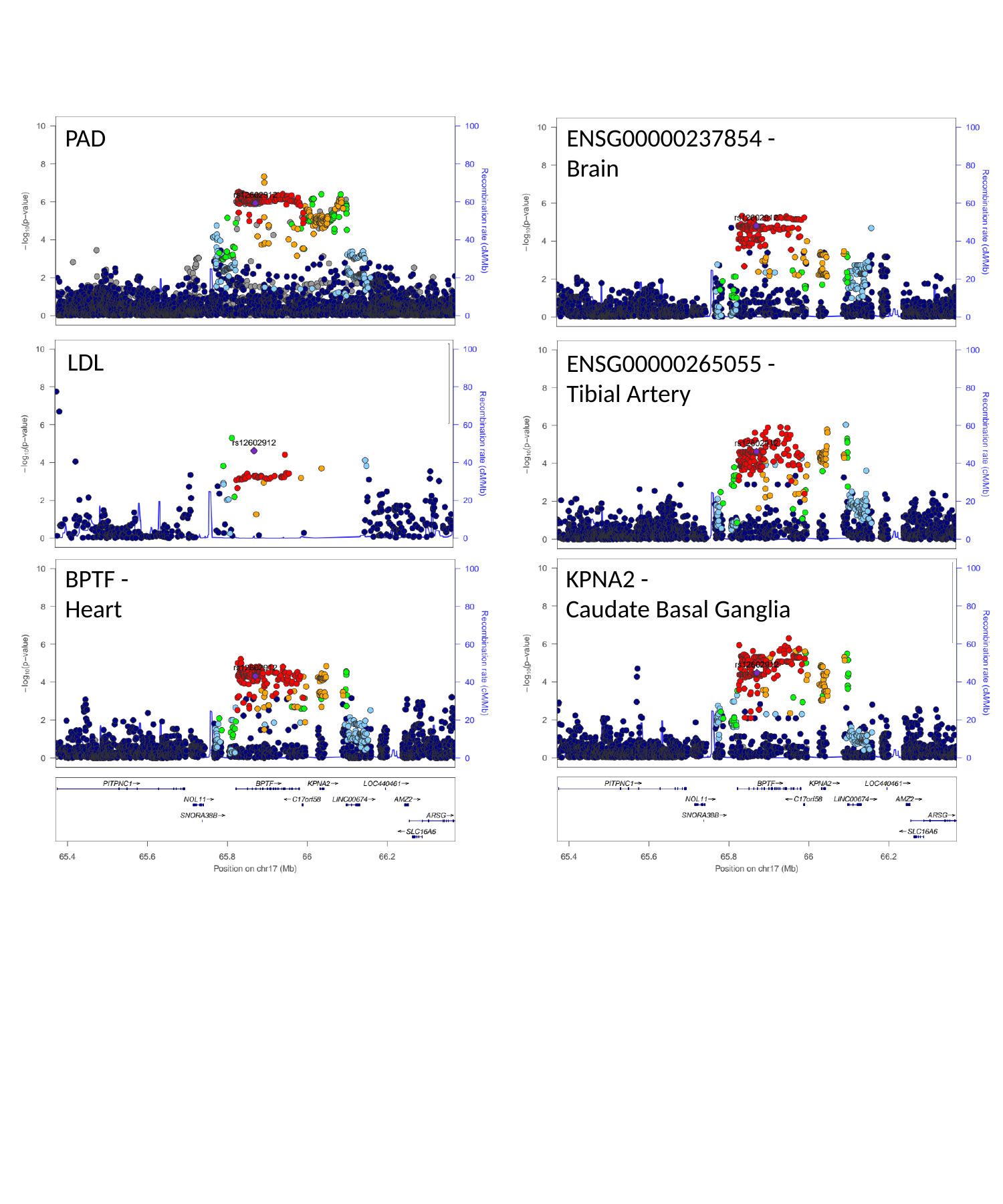

ENSG00000237854 -
Brain
PAD
LDL
ENSG00000265055 -
Tibial Artery
KPNA2 -
Caudate Basal Ganglia
BPTF -
Heart

### Slide 21
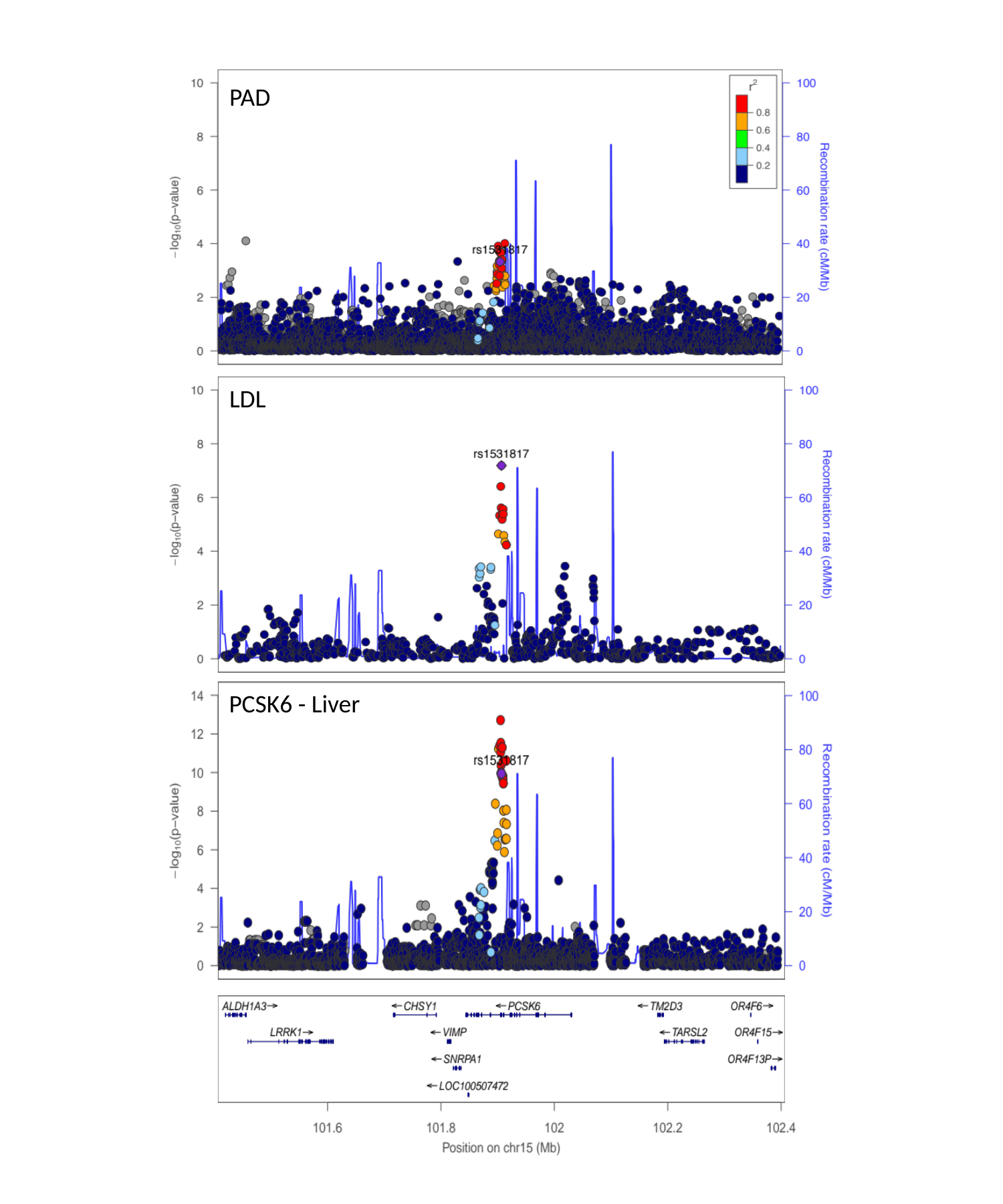

PAD
LDL
PCSK6 - Liver

### Slide 22
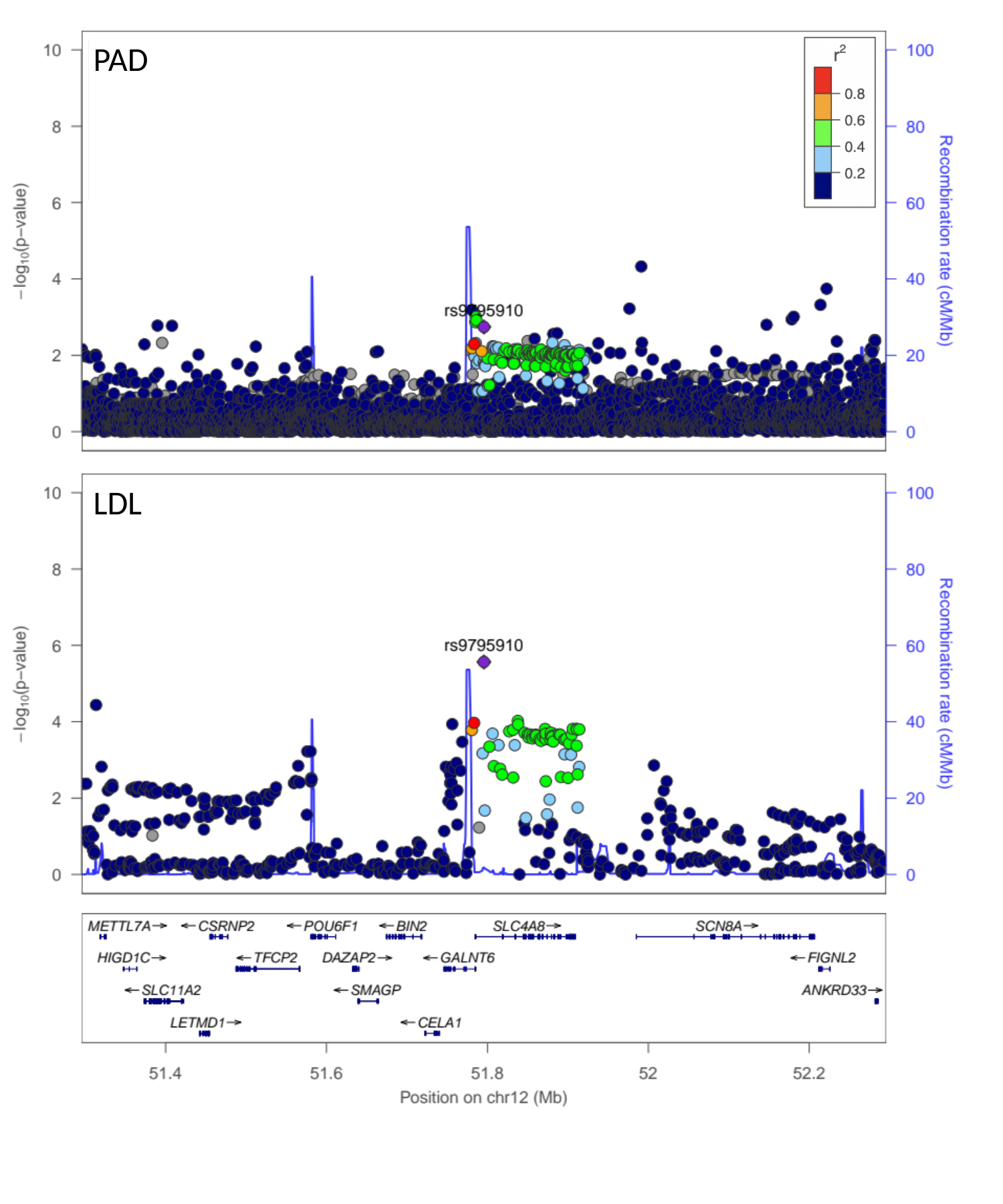

PAD
LDL

### Slide 23
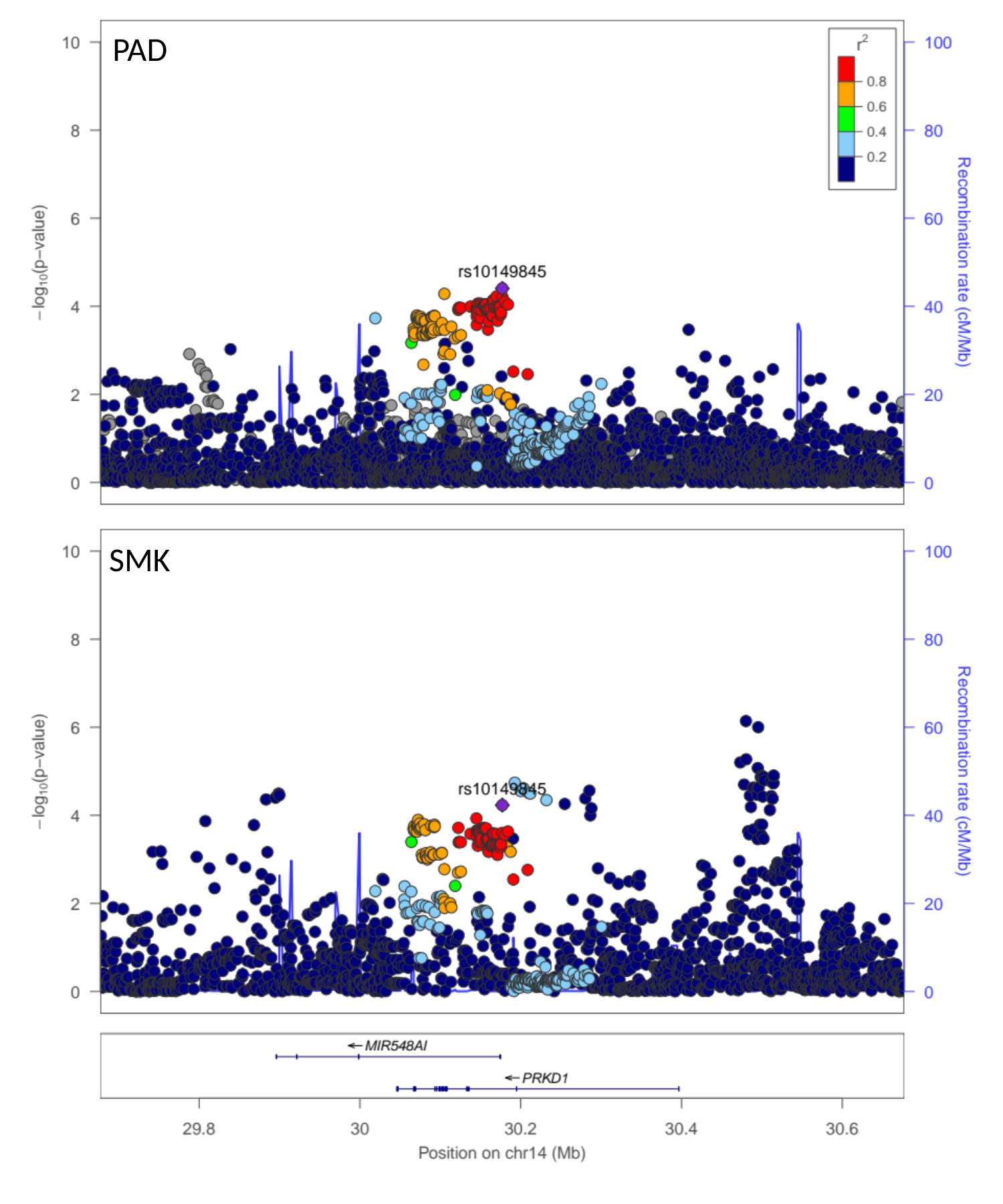

PAD
SMK

### Slide 24
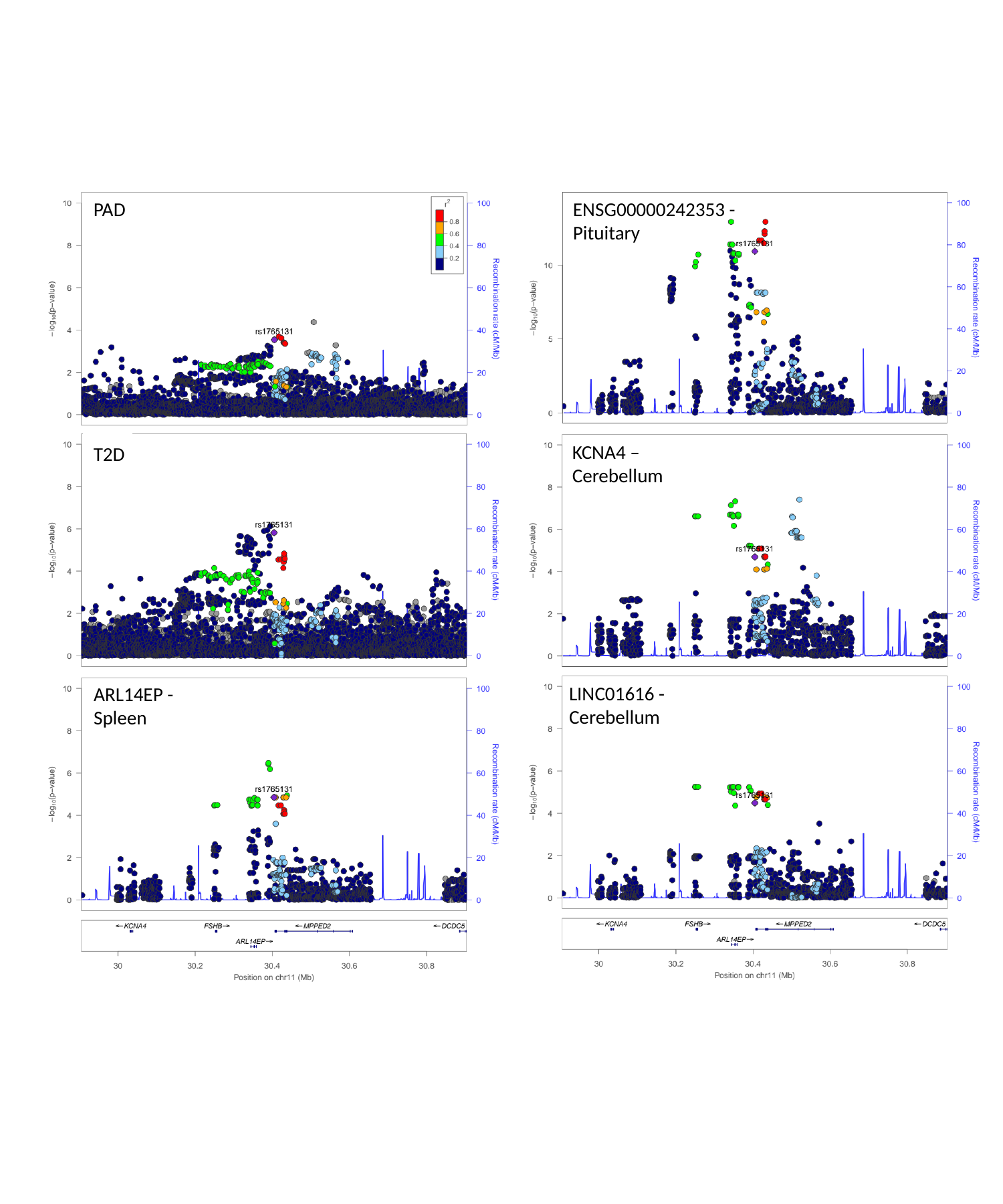

PAD
ENSG00000242353 -
Pituitary
KCNA4 –
Cerebellum
T2D
LINC01616 -
Cerebellum
ARL14EP -
Spleen

### Slide 25
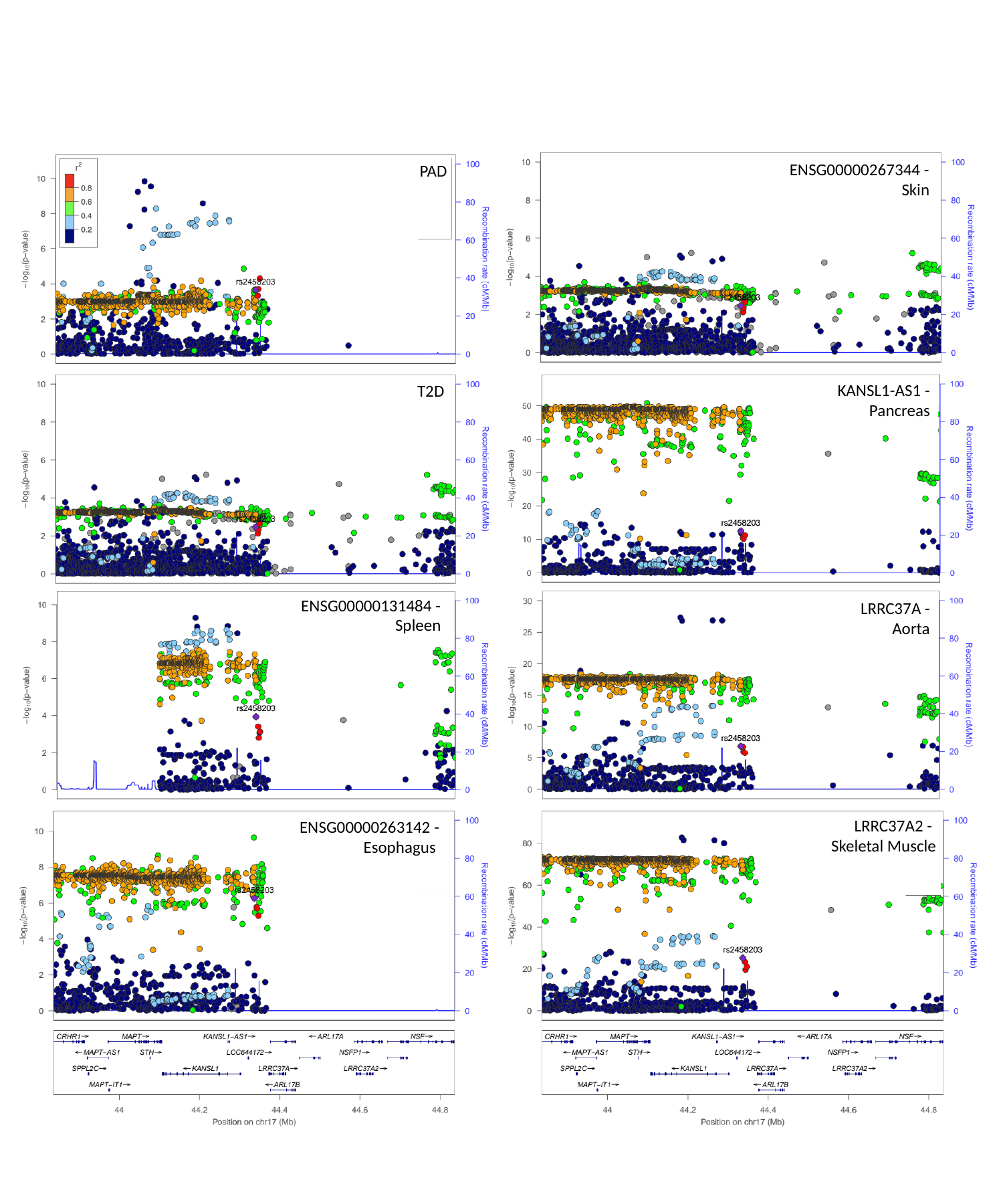

ENSG00000267344 -
Skin
PAD
KANSL1-AS1 -
Pancreas
T2D
ENSG00000131484 -
Spleen
LRRC37A -
Aorta
LRRC37A2 -
Skeletal Muscle
ENSG00000263142 -
Esophagus

### Slide 26
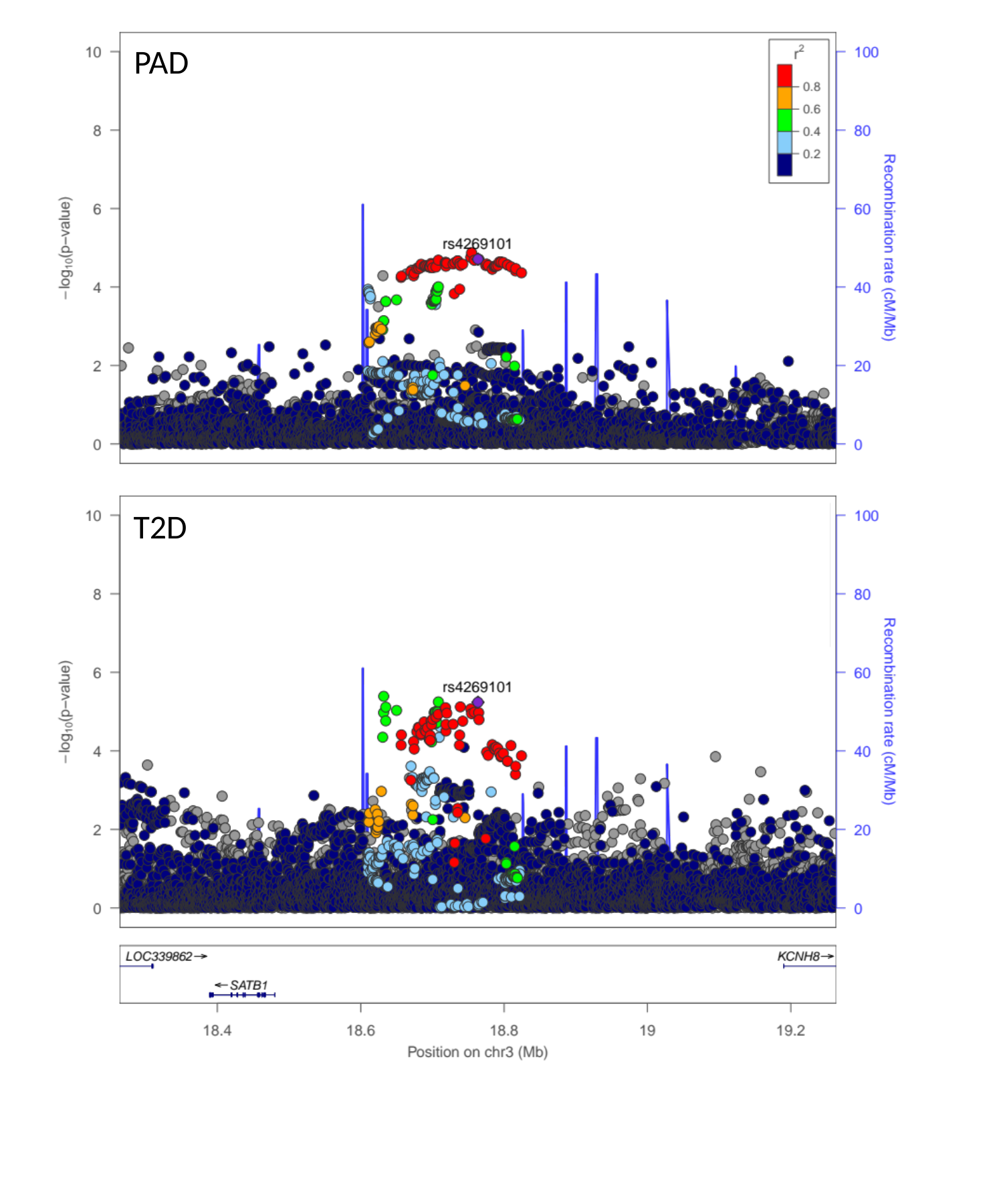

PAD
T2D

### Slide 27
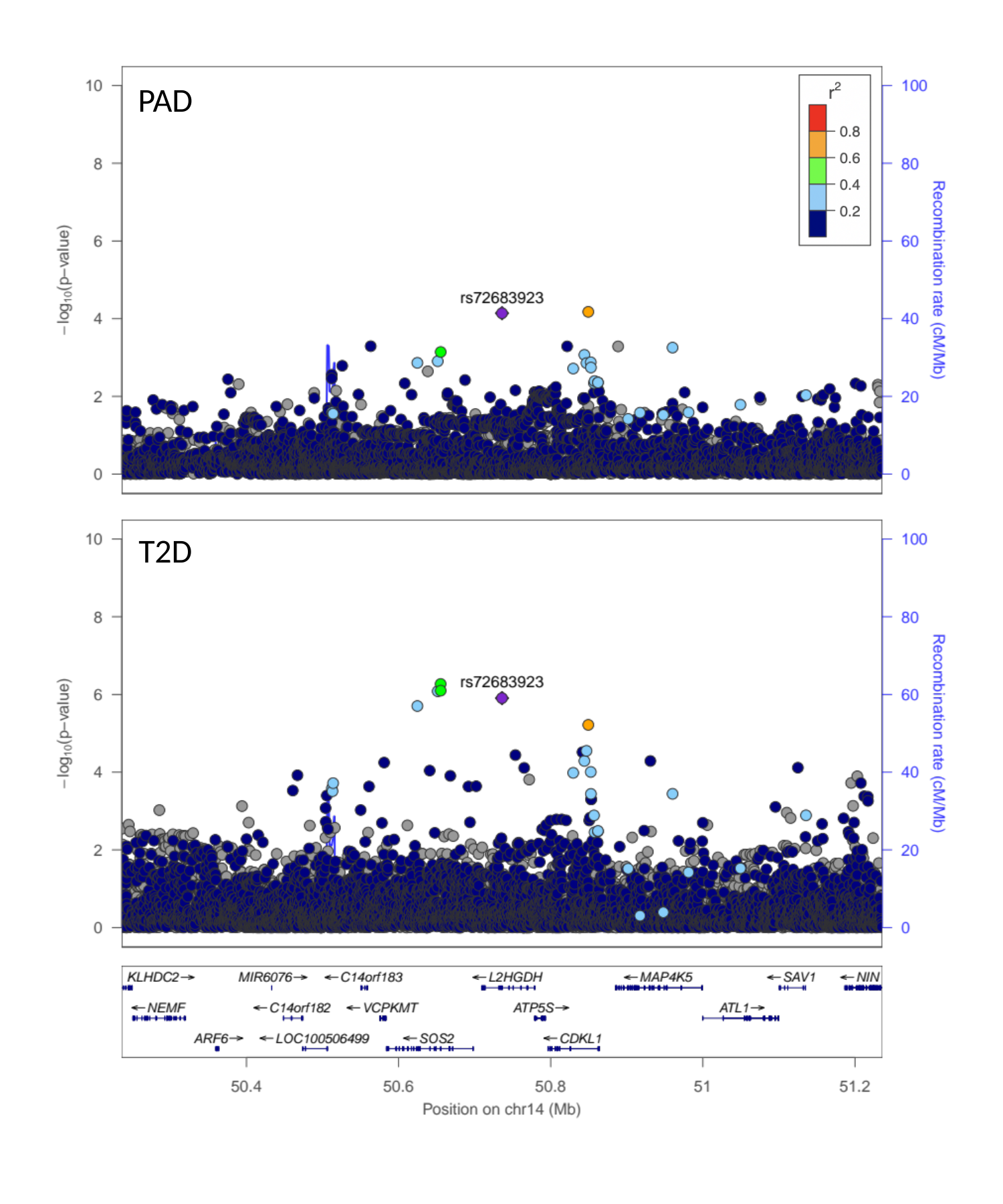

PAD
T2D

### Slide 28
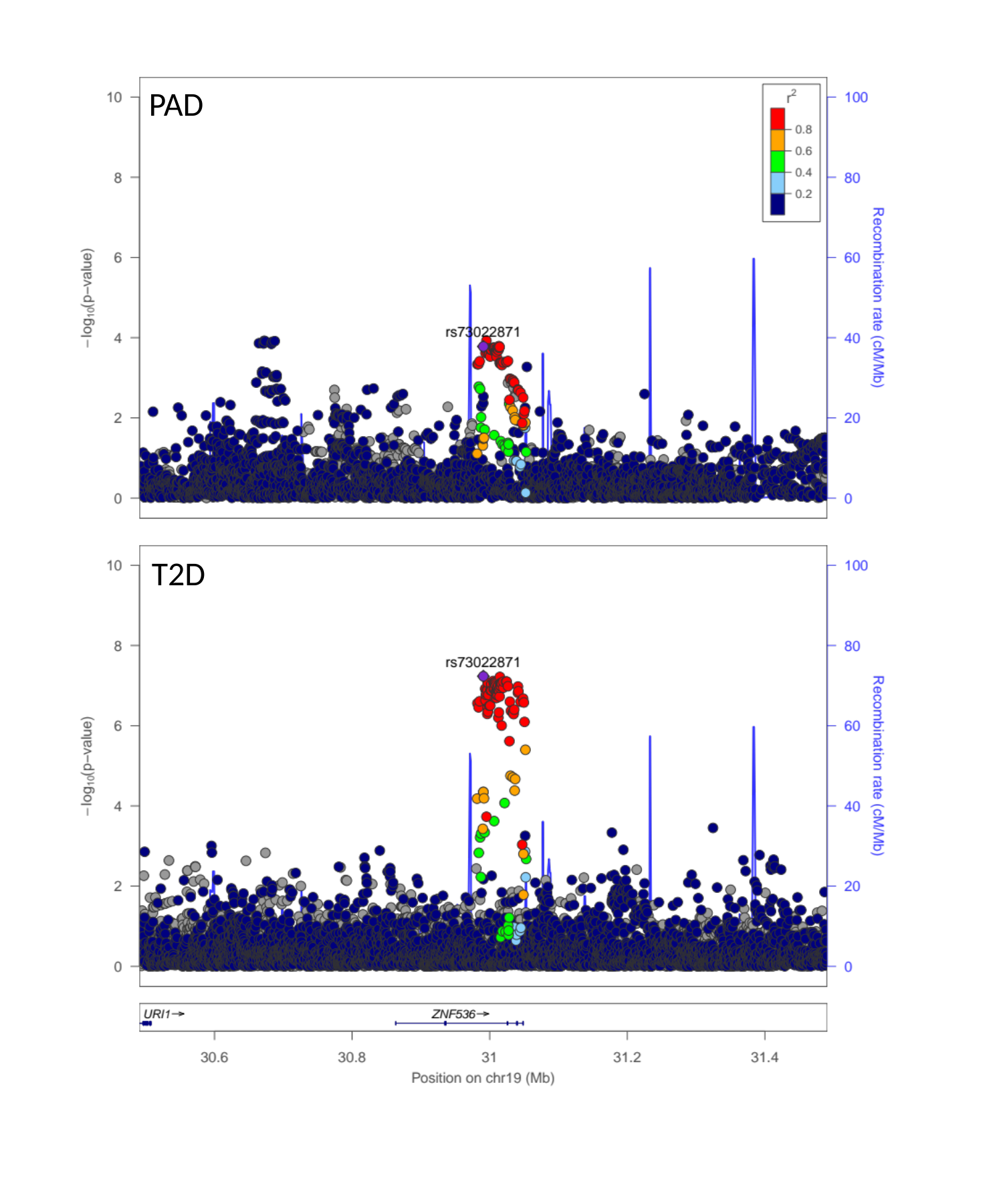

PAD
T2D

### Slide 29
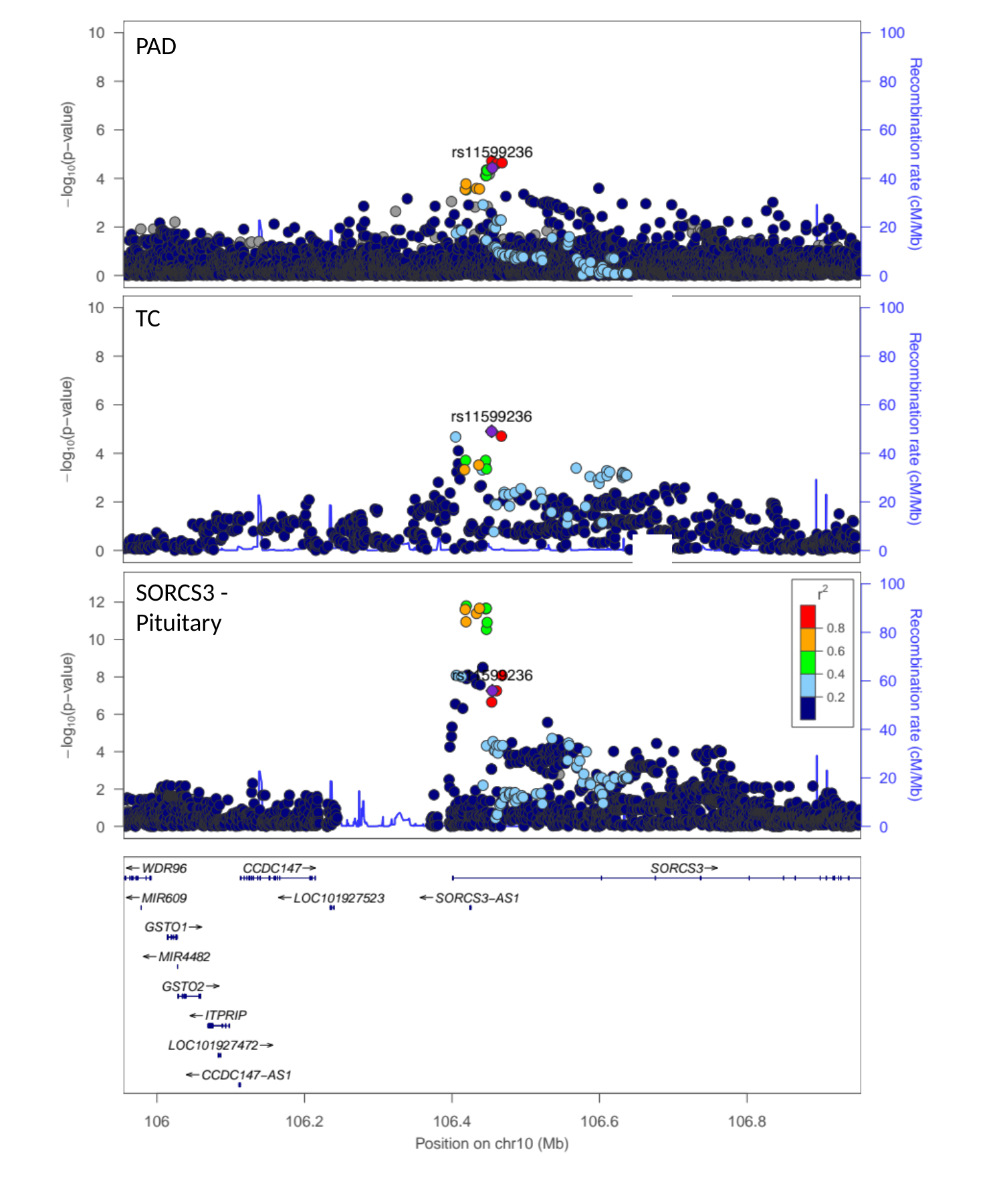

PAD
TC
SORCS3 -
Pituitary

### Slide 30
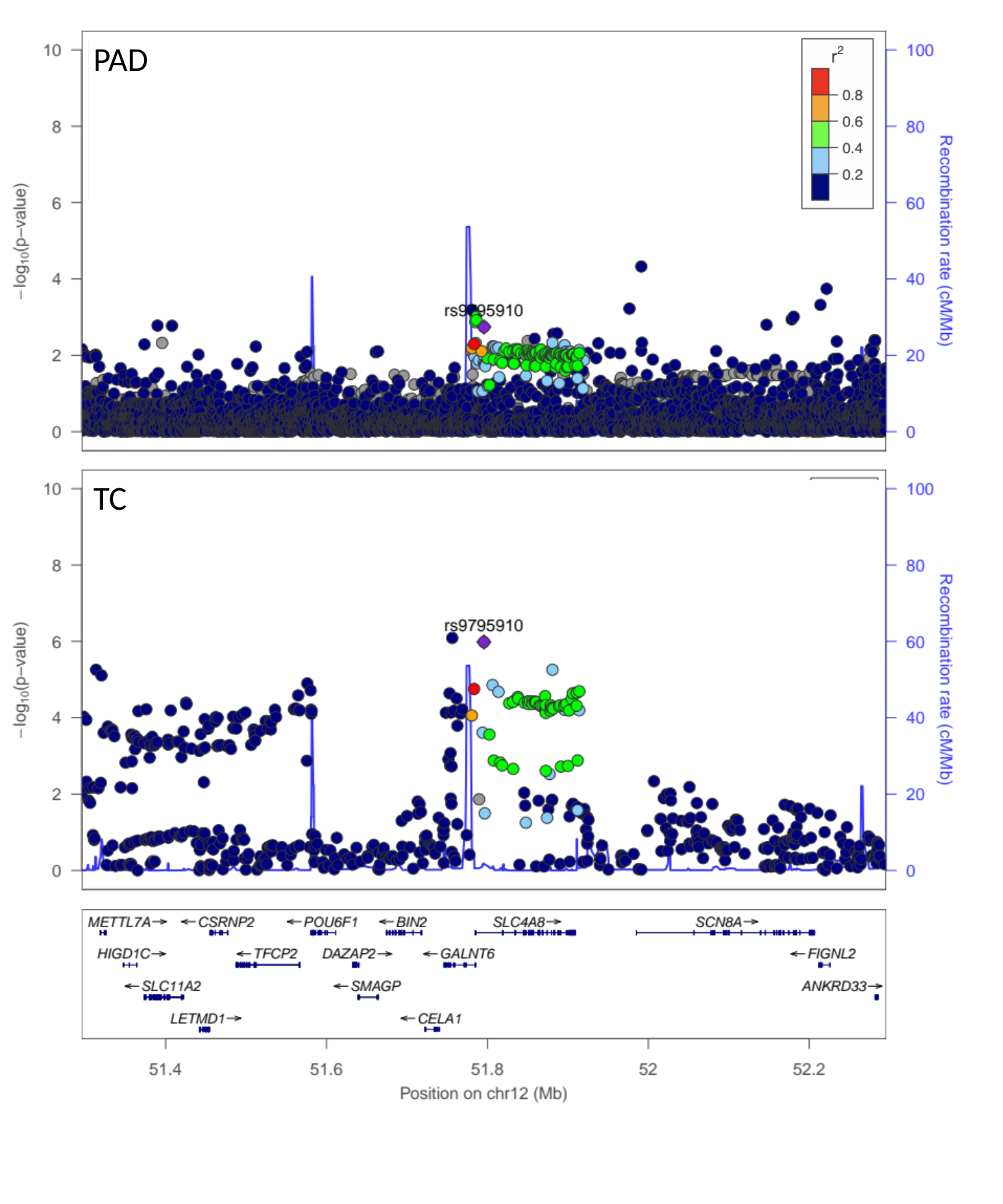

PAD
TC

### Slide 31
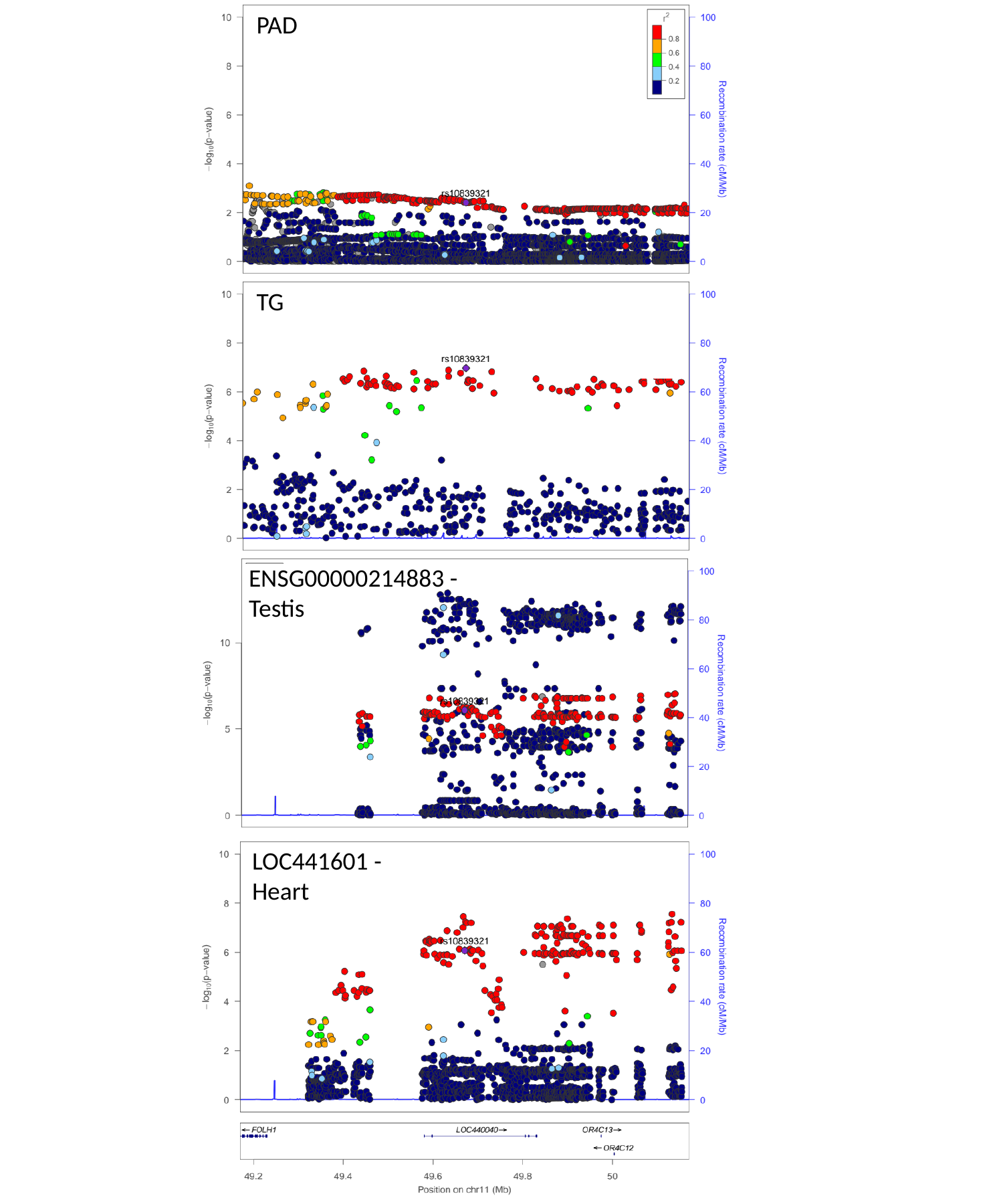

PAD
TG
ENSG00000214883 -
Testis
LOC441601 -
Heart

### Slide 32
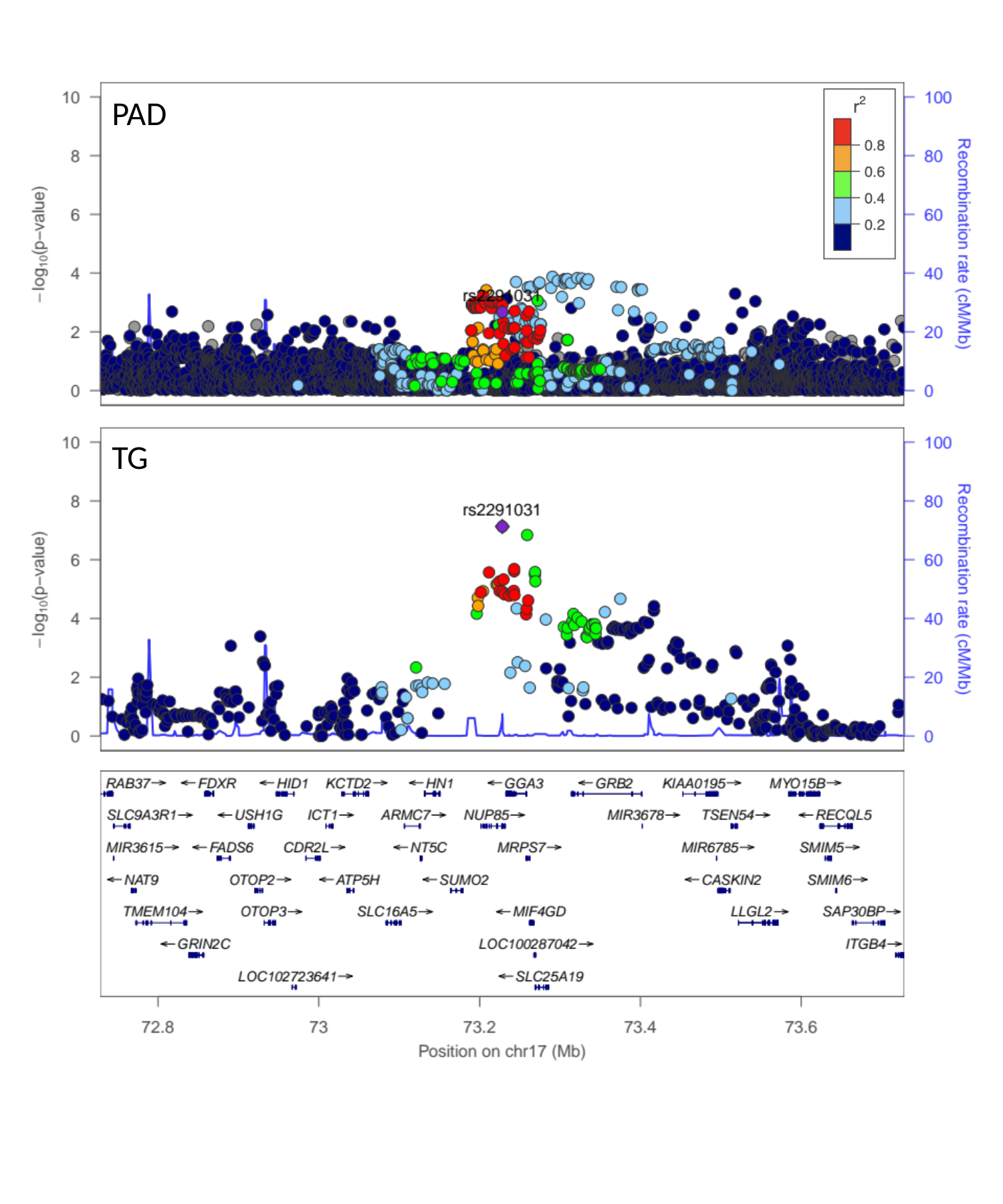

PAD
TG

### Slide 33
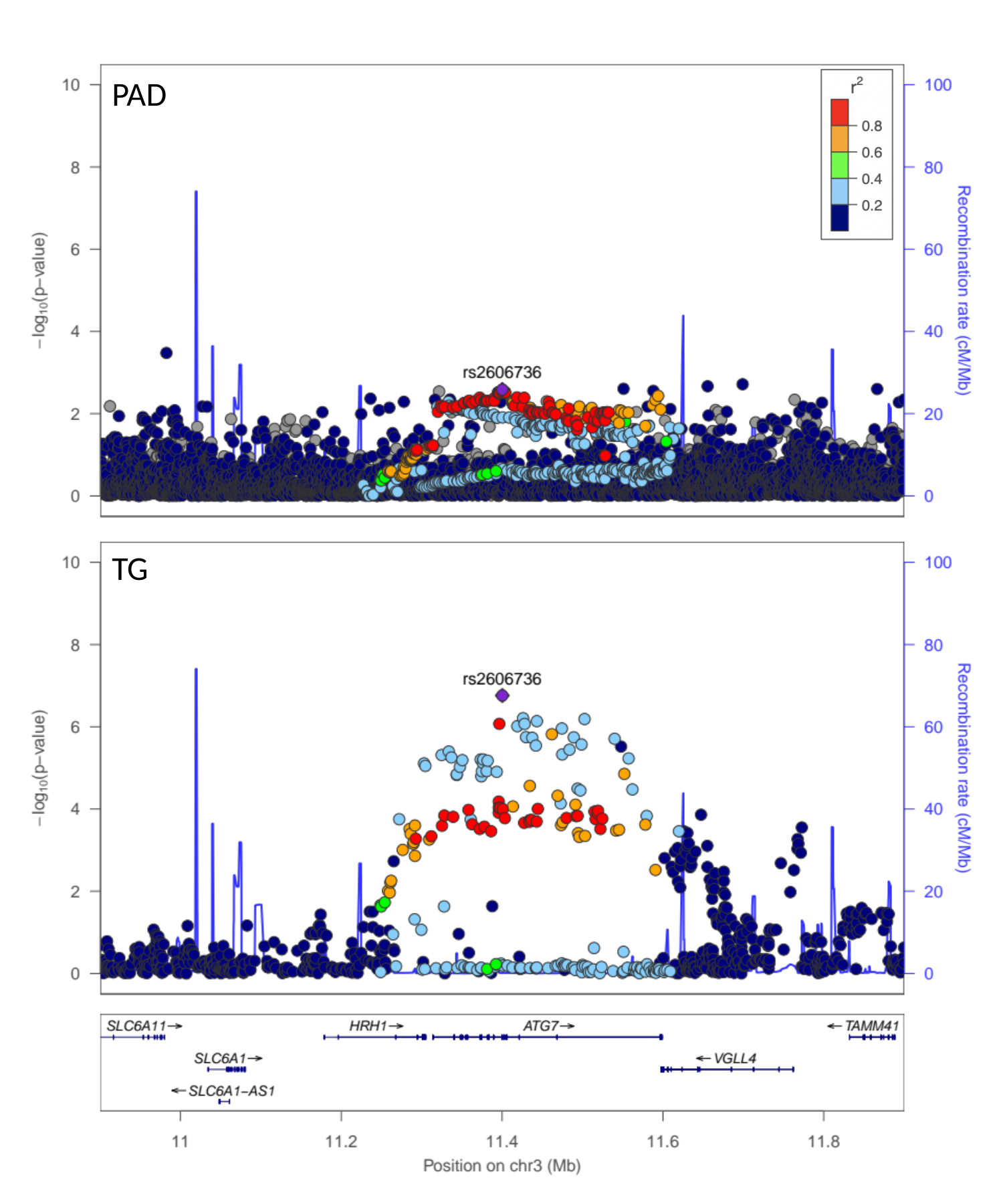

PAD
TG

### Slide 34
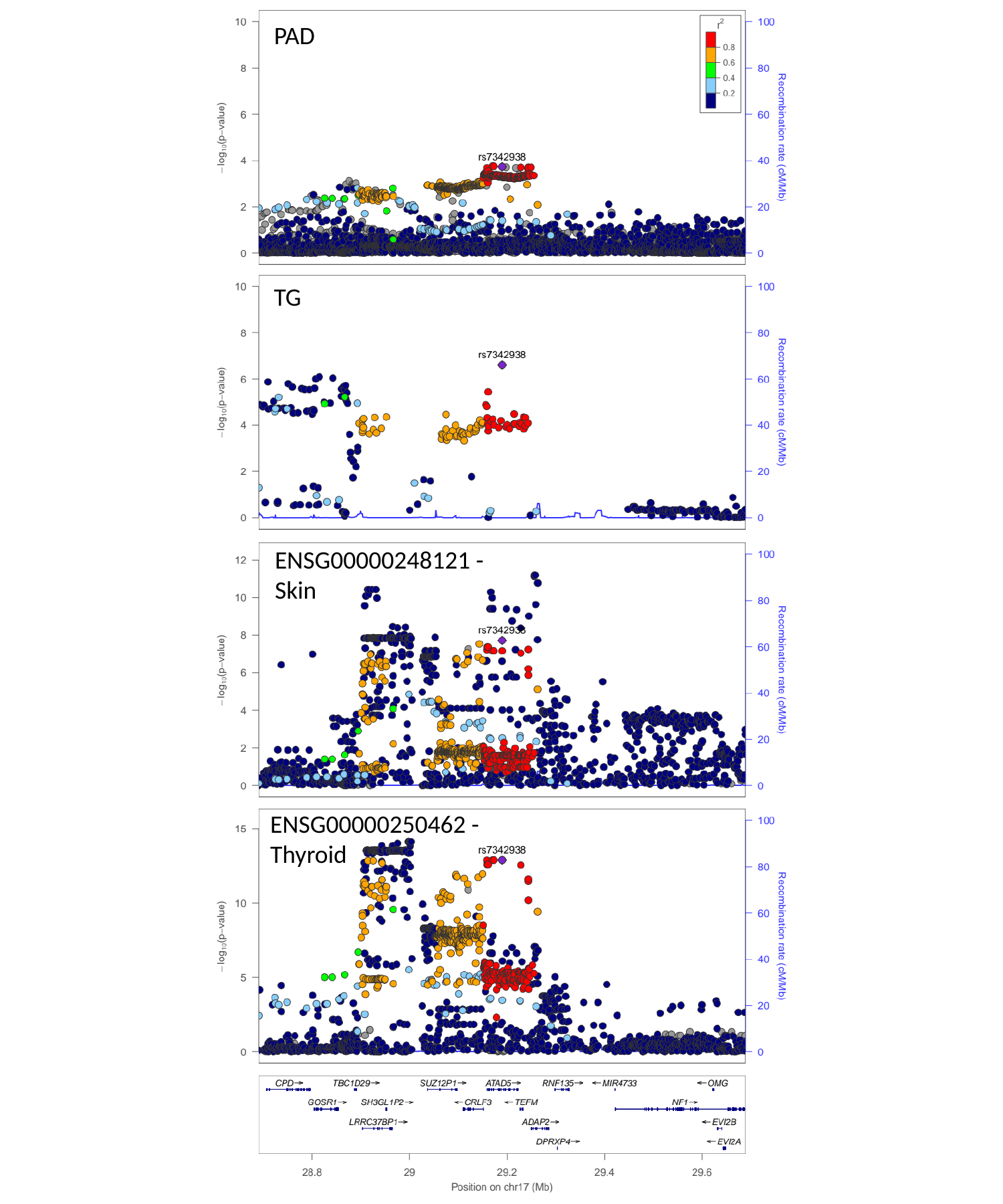

PAD
TG
ENSG00000248121 -
Skin
ENSG00000250462 -
Thyroid

### Slide 35
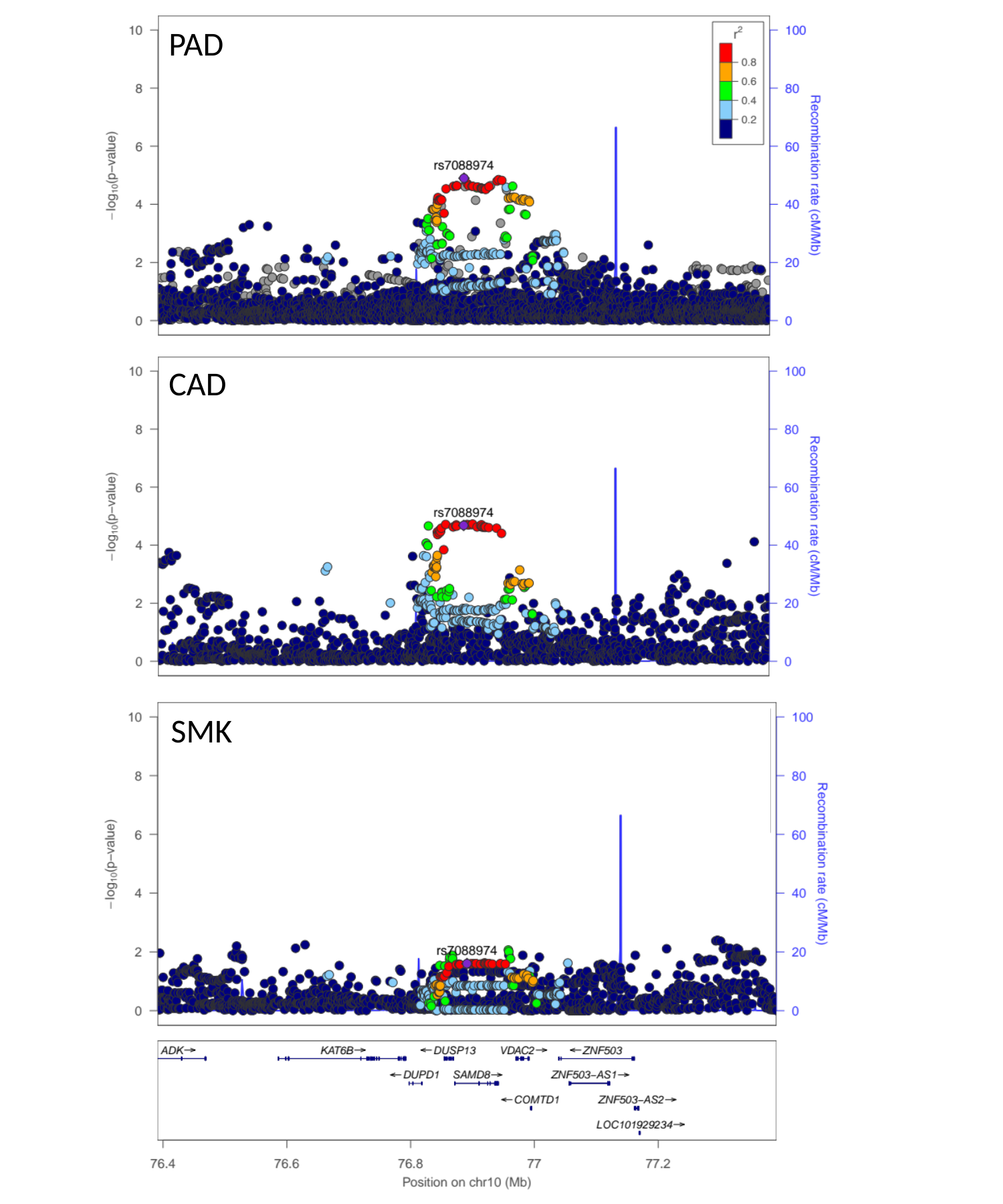

PAD
CAD
SMK

### Slide 36
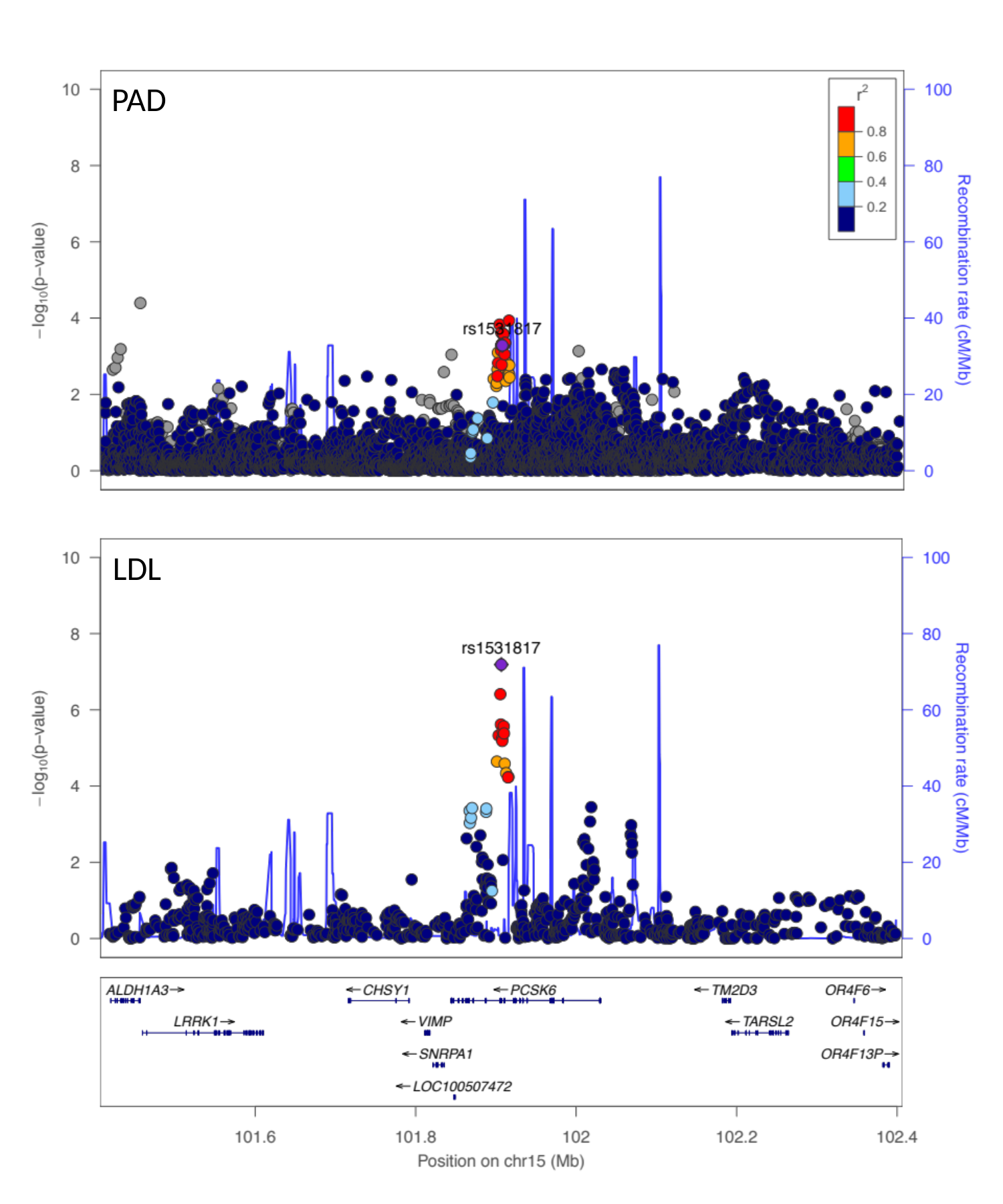

PAD
LDL
