## Supplemental Methods for "Multi-trait GWAS of atherosclerosis detects novel pleiotropic loci"

**N-GWAMA Method**

The N-GWAMA multi-trait GWAS method uses GWAS summary statistics from the univariate trait GWAS to test if there is an association between each SNP with one or more of the traits tested [PMID: 30643256]. N-GWAMA performs this test via a summed weighted Z-score approach. In brief, N-GWAMA uses LD-score regression (LDSC) to estimate the heritability, genetic covariance, and sample overlap of the tested traits [PMID 25642630, 26414676]. Then for each SNP, the univariate Z-scores are weighted by their sample size and estimated heritability. These weights are subsequently added together and that sum is standardized using the estimated variance and covariance of the univariate traits to produce the test statistic Z_k_ (Equation 1). Under the null hypothesis, Z_k_ has a standard normal distribution and thus can easily be used to calculate a p-value for each SNP.

Equation 1:

$$Z_{k}=\frac{\sum_{i=1}^{n} {(w}_{ik}Z_{ik})}{\sqrt{\sum_{i=1}^{n} {(w}_{ik}V)+\sum_{i=1}^{n} \sum_{j=1}^{n} (\sqrt{w_{ik}w_{jk}}C_{i,j})}}$$

**N-GWAMA Multi-trait GWAS Pipeline**

The first step of our pipeline was to filter out SNPs that were not in every input univariate GWAS summary statistics files used and align the alleles of each SNP using *MRbase* [PMID 29846171]. We next ran LDSC on the univariate GWAS files in order to estimate the heritability, genetic covariance, and sample overlap of the traits [PMID 25642630, 26414676]. Using the *MRbase* formatted summary statistic files and the LDSC results, we ran N-GWAMA to perform the multi-trait GWAS on each of the SNPs [PMID 30643256]. From the N-GWAMA results, we defined independent loci at the multi-trait genome-wide significant loci using the PLINK command “--clump-r2 0.2” [PMID 17701901]. Finally, we annotated the resulting multi-trait genome-wide significant loci with the genome-wide significant associations within 500KB or r^2^ > 0.2 from GWAS Catalog. We removed multi-trait genome-wide significant loci from our novel loci list that were annotated with univariate genome-wide significant signals for one or more of the traits involved in the multi-trait GWAS. The code for this pipeline can be found at (Tiffany’s Github).
